# A mechanistic model of curative combination therapy explains lymphoma clinical trial results

**DOI:** 10.1101/2024.06.25.24309486

**Authors:** Amy E. Pomeroy, Adam C. Palmer

**Affiliations:** Department of Pharmacology, Computational Medicine Program, UNC Lineberger Comprehensive Cancer Center, University of North Carolina at Chapel Hill, Chapel Hill, North Carolina, USA

## Abstract

Combinations of chemotherapies are used to treat many cancer types as they elicit higher cure rates and longer responses than single drugs. Several rationales contribute to the efficacy of combinations, including overcoming inter-patient and intra-tumor heterogeneity and improving efficacy through additive or synergistic pharmacological effects. We present a quantitative model that unifies these phenomena to simulate the clinical activity of curative combination therapies. This mechanistic simulation describes kinetics of tumor growth and death in response to treatment and outputs progression-free survival (PFS) distributions in patient populations. We applied this model to first-line combination therapy for Diffuse Large B-Cell Lymphoma, which is cured in most patients by the 5-drug combination RCHOP. This mechanistic model reproduced clinically observed PFS distributions, kinetics of tumor killing measured by circulating tumor DNA, and the adverse prognostic effect of tumor proliferation rate. The outcomes of nine phase 3 trials of new therapies combined with RCHOP were accurately predicted by the model, based on new therapies’ efficacies in trials in patients with relapsed or refractory disease. Finally, we used the model to explore how drug synergy and predictive biomarkers affect the chance of success of randomized trials. These findings show that curative combination therapies can be understood in quantitative and kinetic detail, and that predictive simulations can be used to aid the design of new treatment regimens and clinical trials in curative-intent settings.

**SIGNIFICANCE:** A novel model that incorporates pharmacological interactions in the presence of inter-patient and intra-tumor heterogeneity explains and predicts combination clinical trial outcomes of curative regimes used to treat Diffuse Large B-cell lymphoma. This model can be used to understand and inform optimal design of drug combinations and clinical trials.

## INTRODUCTION

Combination chemotherapy is a foundation of treatments with curative intent for advanced cancers such as lymphomas and leukemias. The original rationale for combination therapy was to overcome tumor heterogeneity (1), which occurs both within patients (‘intra-tumor’) and between patients (‘inter-patient’). Theoretical modeling and analysis of each of these phenomena individually have contributed to our knowledge about responses to cancer treatment. Currently, there is an unmet need to combine both sources of heterogeneity into one framework, as both intra-patient and inter-tumor heterogeneity are key influences on the efficacy of cancer treatments, especially combination therapy. The relevance of intra-tumor heterogeneity to combination therapy was described in 1952 by Lloyd Law, who proposed that although single drugs are resisted by a fraction of cells in a tumor, drug combinations may be resisted by only a fraction of a fraction of cells, which might be small enough to eradicate a cancer (2). The role of inter-patient heterogeneity was observed in 1962 by the Acute Leukemia Group B, who found that combination therapy can increase the percentage of patients who respond, because some patients’ cancers respond to one drug and some patients’ cancers respond to a different drug (3). Recently we experimentally confirmed Law’s hypothesis in the context of Diffuse Large B-Cell Lymphoma cultures treated with the curative RCHOP regimen (4). We also found that inter-patient heterogeneity explains much of the clinical efficacy of modern combination therapies for incurable cancers (5,6). These prior works investigated intra-tumor and inter-patient heterogeneity separately, but both phenomena are relevant to cancers in human populations.

To understand the joint effect of inter-patient and intra-tumor heterogeneity, we recently described simple models which propose that because single drugs have variable efficacy among patients, drug combinations can be viewed as a sum of variable effects in each patient (7). Applied to fractional tumor cell kill, this concept explains historical improvements in response and cure rates in pediatric acute lymphocytic leukemia (7). Applied to Progression-Free Survival (PFS) data, this concept explains the PFS benefits of most drug combinations to have received FDA-approval for advanced cancers in the past 25 years (8). These phenomenological models have conceptual and practical value, but a mechanistically explicit model, grounded in pharmacology and tumor biology, is desirable to better understand how combination therapies sometimes succeed and sometimes fail to overcome tumor heterogeneity. In particular, understanding curative combinations will require models that explicitly consider dose responses, treatment schedules, and tumor kinetics.

To develop a mechanistic model of tumor responses to drug combinations, here we begin by considering the seminal work of Skipper, Schabel, and Wilcox, who showed in mice with leukemia that a dose of chemotherapy kills a constant fraction of cancer cells, known as the ‘fractional kill hypothesis’ (9). An extrapolation of this hypothesis has become a textbook figure, showing a constant fraction of tumor population killed by each cycle of therapy (1,10) (**Figure 1A**). However, Skipper’s experiments did not suggest that subsequent doses produce the same cytotoxic effect as the first, and this extrapolation leads to a false conclusion: that any initially responsive tumor could be cured with enough cycles of chemotherapy. Past models have resolved this deficit by including drug-resistant subpopulations of cancer cells, which are selected by therapy and cause relapse (**Figure 1A**). Variations on this approach have produced many insights into how the evolution of drug resistance can inform regimen design (11–14). Describing cancer cells simply as drug ‘sensitive’ or ‘resistant’ may be appropriate for cases such as BCR-ABL inhibition where resistance is chiefly due to binding site mutations (15). However, this binary model is generally over-simplistic, being unable to capture the spectrum of treatment responsiveness in human populations, or to explain how it could be that some cancers are curable even though treatment always selects for resistant subpopulations. Here we show that by moving beyond a binary view of drug resistance, modeling intra-tumor and inter-patient heterogeneity as distributions of drug sensitivity phenotypes can reproduce survival distributions in curative settings.

**Figure 1.**
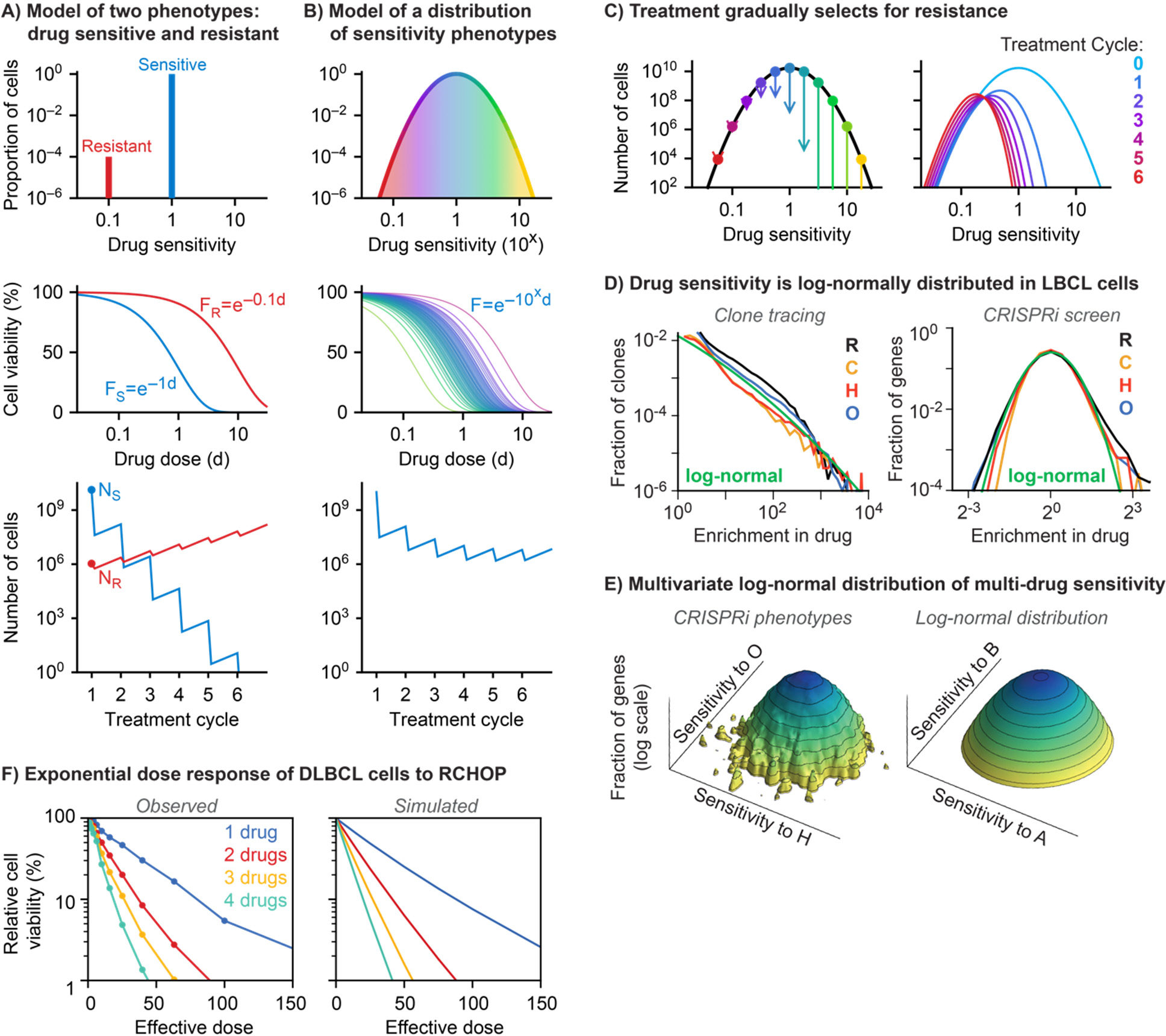
Modeling intra-tumor heterogeneity in drug sensitivity as a distribution. (A) When tumor heterogeneity is modeled as two phenotypes – sensitive and resistant – with dose response functions F_S_ and F_R_ respectively, a constant fraction of sensitive cells is killed by each cycle of treatment, but the drug-resistant subpopulation survives therapy and causes disease relapse**. (B)** When tumor heterogeneity modeled as a continuous distribution of sensitivity phenotypes, where cell as a distinct dose response depending on its drug sensitivity, a large fraction of cells is killed by the first cycle of treatment and decreasing fractions of cells killed by each subsequent cycle. **(C)** In this model, the distribution of drug sensitives in the tumor shifts towards resistance (lower drug sensitivity) with each cycle of treatment as highly sensitive cells are killed and resistant cells survive. **(D)** Both the distribution of clone enrichment in cells treated with either rituximab (R), cyclophosphamide (C), doxorubicin (H), or vincristine (O) and the distribution of the effect of CRISPR inhibition (CRISPRi) on the sensitivity of DLBCL cells to each drug follow a log-normal distribution. **(E)** Two-dimensional distribution of the effect of CRISPRi on sensitivity of cells to doxorubicin and vincristine follow a to a two-dimensional log-normal distribution. **(F)** Experimentally observed dose response curves of DLBCL cells to treatment with combinations of either one, two, three, or four drugs in RCHO compared to model simulations using exponential dose response curves.

Distributions of phenotypes are routine in physiology and pharmacology, and are an expected consequence of the central limit theorem whenever there are many sources of variation (16). This property has long been exploited by ‘mixed effect’ models of population pharmacokinetics (pop-PK), which use parameter distributions to describe patient-to-patient heterogeneity in pharmacokinetic rates (17). This approach does not need to explicitly model each mechanism of variation; instead, it fits a magnitude of inter-patient variance to observed pharmacokinetic variation, implicitly accounting for biological variation even when underlying mechanisms are unknown. This approach could be particularly helpful to model drug resistance in cancer because the causes of chemotherapeutic resistance are diverse and poorly understood. Fitting to human distributions of treatment effect can capture the *net* impact of many sources of drug response heterogeneity, including genetics, epigenetics, tumor microenvironment, drug bioavailability, and others (without needing to settle debates about which is most important). Mixed-effect models have been used before to model inter-patient variance for *in silico* clinical trials (18). In our population-tumor kinetics (pop-TK) model, we represent both inter-patient and intra-tumor heterogeneity as distributions, enabling a detailed simulation of chemotherapy response that accounts for variability at both patient and cellular levels.

We apply the ‘pop-TK’ model to the most common blood cancer, Diffuse Large B-Cell Lymphoma (DLBCL). The potential to cure DLBCL was demonstrated in the 1970s by DeVita and colleagues, leading to the establishment of a four-drug combination of cyclophosphamide, doxorubicin, vincristine, and prednisone, abbreviated as CHOP (19,20). Efficacy was further improved in 2002 by the addition of Rituximab, making the 5-drug RCHOP regimen (21). Many subsequent trials failed to improve upon RCHOP (22–31), until recently the Pola-R-CHP regimen become in over 20 years to significantly improve progression-free survival in previously untreated DLBCL (24). The history of combination therapy trials in DLBCL raises an important question: why do some new combination therapies improve cure rate while others do not? Here we use a mechanistic model to understand how cures are achieved by current regimens, with the goal of informing the design of new regimens with curative intent.

## RESULTS

### Modeling tumor response to cycles of combination therapy

Our model of heterogeneous cancers treated with drug combinations is explained here by building up in complexity. We begin with a simple, familiar framework of one drug acting on one patient, whose cancer contains drug-sensitive and resistant cells with initial populations *N_S_* and *N_R_*, where fractions *F_S_*(*d*) and *F_R_*(*d*) respectively survive one cycle of treatment with drug at dose *d* (**Figure 1A**). The total number of cells that survive treatment will be *N_S_F_S_*(*d*) + *N_R_F_R_*(*d*). Applying this model to repeated cycles of therapy with growth between cycles produces Darwinian selection for drug resistance (**Figure 1A**). If heterogeneity in drug sensitivity is described as a series of many states rather than two extremes, the total population after treatment can be described as a sum ∑_*i*_ *N_i_F_i_*(*d*). It follows then, that a sum of many states can be described as an integral over a distribution of drug sensitivity levels *x*: ∫ *N*(*x*)*F*(*x*, *d*)*dx* (**Figure 1B**). In this view, the consequence of intra-tumor heterogeneity is that the cytotoxic effect of chemotherapy is described by an ensemble of dose-response functions, with some cells more sensitive than average and some more resistant than average. Greater or lesser initial drug sensitivity is defined by the initial center of the drug sensitivity distribution. Treatment preferentially kills cells on the more drug-sensitive side of the distribution, such that repeated cycles of therapy progressively shifts a tumor population toward resistance (**Figure 1C**), reproducing a common clinical scenario of initial drug response followed by relapse. Cancer cells can resist chemotherapy for a multitude of reasons. Here, rather than modeling specific mechanisms, all physiologically relevant causes of resistance are viewed as reasons why some (perhaps many) tumor cells lie on the resistant side of the distribution. Darwinian selection therefore emerges from this model in a mechanism-agnostic manner.

This model extends to combination therapy by considering multiple dimensions of drug sensitivity, and multi-drug response functions. For two drugs X and Y, we describe a tumor cell population by a two-dimensional distribution of drug sensitivities *x* and *y*, *N*(*x*, *y*), and a function describing the fractional survival of cells with sensitivities *x* and *y* to dose *d_X_* of drug X and *d_Y_* of drug Y, *F*(*x*, *y*, *d_X_*, *d_Y_*). The total number of cells surviving multi-drug treatment is given by a double integral:

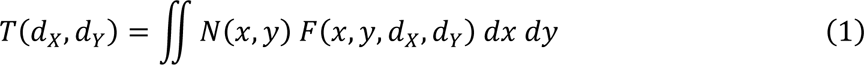

Cross-resistance between drugs manifests as a correlation in the sensitivity distribution *N*(*x*, *y*), and the multi-drug response function *F* can include synergistic, additive, or antagonistic drug interactions (**Methods**). This framework similarly expands to any number of drugs.

This approach has separated tumor response to chemotherapy into two parts: the distribution of tumor cells’ drug sensitivities *N*, and their dose response function *F*. These functions will depend on the drugs and cancer type of interest and are both measurable. Drug sensitivity of cancer cells *in vivo* depends on many factors, including genetics, epigenetics, microenvironment, pharmacokinetics and more. When a quantitative phenotype is the product of many factors, its population distribution can usually be approximated by a log-normal distribution, as a consequence of the central limit theorem (**Figure S1A-B**). Indeed, for DLBCL sensitivity to drugs in RCHOP, we re-analyzed published high complexity clone-tracing experiments and genome-wide CRISPR inhibition screens under therapy (32), and observed that drug sensitivity phenotypes in DLBCL cells are approximately log-normally distributed (**Figure 1D**). Further, considering sensitivity to multiple drugs in RCHOP, experimental data closely resembles partly correlated multivariate log-normal distributions, and specifies magnitude of correlation for RCHOP (**Figures 1E** and **S1C-D**). Determinants of chemosensitivity and tumor heterogeneity are surely more complex *in vivo* than *in vitro*, which strengthens rather than weakens the rationale for describing drug sensitivity as a distribution rather than simple extremes. This approach is helpfully agnostic to our limited knowledge of *in vivo* determinants of chemosensitivity.

Turning to dose response *F*, like most cytotoxic chemotherapies the drugs in RCHOP exhibit approximately exponential dose response functions, both as single drugs and when combined (1,32) (**Figure 1F**). Although some pairs of drugs in RCHOP interact antagonistically, response to the full five-drug combination is observed to be additive (non-interacting) according to both Loewe’s and Bliss’ definitions of synergy (32). The effect of differing drug sensitivity is modeled using a previously reported approach of rescaling drug concentration, such that drug resistant cells behave as though exposed to less drug (33,34). Rescaling drug concentration is specifically true for resistance mechanisms that decrease target binding (e.g. efflux pumps, binding site mutations, drug degradation, loss of pro-drug activation), and is also observed to approximate the effects of diverse resistance mechanisms in settings ranging from anti-bacterial drugs to DLBCL chemotherapy (32).

Assembling these descriptions of *N* and *F* yields the following integral for the total number of cancer cells (*T*) after *c* cycles of a single drug:

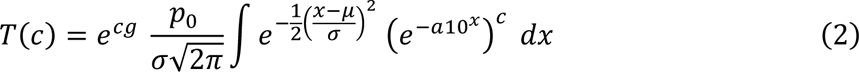

Here the first term after the integral is a normal distribution of *x* which denotes log(drug sensitivity), where *μ* and *σ* are its average and standard deviation; 10^x^ is drug sensitivity. The dose response function is given by (*e*^-a10^x^^)^c^, where *c* is the number of cycles and *a* is the dose of drug, which is rescaled by 10^x^ to account for differing levels of drug sensitivity in a heterogeneous tumor. The integral is multiplied by a normalization constant such that initial number of cancer cells equals *p*_0_. The term *e*^cg^ describes tumor cell growth with rate *g* between treatment cycles. As noted, this framework generalizes to combination therapy with *n* drugs by an *n*-dimensional integral over a multivariate distribution (methods). For combinations of 1 through 5 drugs this model reproduces experimentally measured dose responses of DLBCL cultures treated with combinations of the drugs in RCHOP (Figure 1F). With a model of intra-tumor heterogeneity within one patient, we next consider inter-patient heterogeneity in the setting of a clinical trial.

A clinical trial is modeled as a virtual cohort of heterogeneous patients. Thus, the equation above is applied to each of many simulated patients having their own values for average drug sensitivity (*μ*), initial number of cancer cells (*p*_0_), and tumor growth rate (*g*). Tumor growth rates and initial population sizes are sampled from clinically measured distributions (35,36), and drug sensitivity distributions are fitted to reproduce clinically measured Progression-Free Survival (PFS) distributions. Variation among patients in average drug sensitivity means, for example, that some patients’ entire cancer cell populations are refractory to treatment (**Figure 2A**, red), whereas other patients may be exceptional responders with highly drug sensitive cancer cells (**Figure 2A**, yellow). Various treatment schedules can be simulated, producing trajectories of tumor cell population over time, which determine whether a virtual patient is cured, or if not, when they experience disease progression (**Figure 2B**). A cohort of outcomes yields a Kaplan-Meier plot of PFS (**Figure 2C**). Three parameters chiefly influence drug activity: average drug sensitivity across all patients, inter-patient variance (*σ_patients_*), and intra-tumor or cell-to-cell variance (*σ_cells_*); these affect PFS distributions in distinct ways and can therefore be fitted to clinical trial results (**Figure S2**).

**Figure 2.**
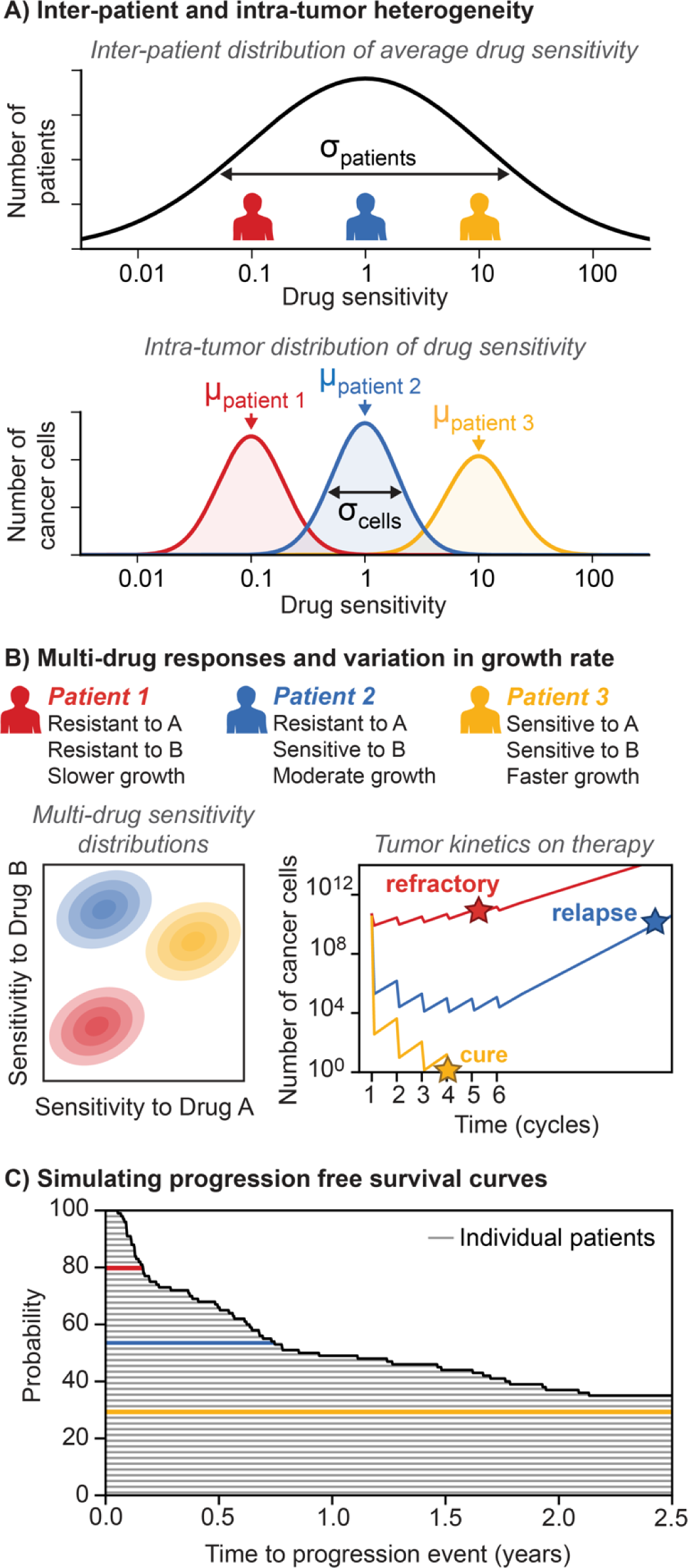
Incorporating inter-patient heterogeneity to simulate clinical outcomes in populations. (A) The drug sensitivity of cohorts of patients is simulated by sampling each patient’s drug sensitivity from a log-normal distribution with mean, *μ_patients_*, and inter-patient heterogeneity, *σ_patients_*. Each individual patient then has a unique average sensitivity, *μ_patient_* _N_, and an intra-tumor heterogeneity, *σ_cells_*, which is the same for each patient. **(B)** When simulating combinations of multiple drugs, each patient has multidimensional sensitivity distribution, and tumor trajectory for each patient can be calculated based on their drug sensitivities, growth rate, and initial tumor size. Representative trajectories for three patients are shown. **(C)** Progression times calculated from tumor trajectories are used to generate progression free survival (PFS) distributions for cohorts of heterogenous patients.

### Explaining key features of DLBCL clinical response

Each drug in RCHOP has a distinct mechanism of action and is therefore modelled as acting on a distinct axis of drug sensitivity (**Figure 2B**). We calibrated our model on PFS distributions from the CHOP and RCHOP arms of the RICOVER-60 trial (37). Specifically, the Metropolis-Hastings algorithm fitted six parameters (mean sensitivity, inter-patient and intra-tumor heterogeneity, inter-patient and intra-tumor cross-resistance, and average tumor growth rate) (**Figure S3**), with the remaining model parameters directly set by clinical measurements (**Supplemental Table S1**). The algorithm returned an ensemble of parameters sets that fall within the 95% confidence interval of observed PFS distributions and reproduce the benefit of Rituximab (**Figure 2A**). The primary source of parameter flexibility was a trade-off between intra-tumor heterogeneity and average drug sensitivity, because greater heterogeneity entails more resistant cells. The parameter set closest to the average of the ensemble was selected as a representative set for subsequent analyses (**Figure S4**).

Fitting only to the PFS distribution of RCHOP yielded a model that also reproduced kinetic features of DLBCL treatment. Liquid biopsies to measure circulating tumor DNA (ctDNA) have recently enabled highly sensitive monitoring of lymphoma response during RCHOP treatment (**Figure 2B**). The observed distribution of ctDNA changes in patients after their first cycle of RCHOP was qualitatively similar to its simulated equivalent over 5 orders of magnitude, as was the proportion of patients with undetectable disease after the first cycle (observed 28%; simulated 33%), indicating that the model reproduces the initial decline of tumor population (**Figure 2B**). Further, Ki67 staining is a marker of tumor cell proliferation and an adverse prognostic factor in DLBCL (38–43). We stratified simulated patients by their tumor proliferation rate (a parameter with inter-patient variation) and found it to be an adverse prognostic factor in the model (Hazard Ratio = 1.3, p<0.01), similar to clinical observations (**Figure 2C**). Together, these results demonstrate that fitting the pop-TK model to PFS also reproduces tumor death kinetics and prognostic effect of tumor growth rate.

### Predicting DLBCL combination therapy trials based on monotherapy outcomes

Since the development of RCHOP, many phase 3 trials tested new drugs in combinations for first-line DLBCL, nearly all of which were preceded by a trial of the new drug in relapsed or refractory (R/R) DLBCL (22–31,44) (**Table S2**). Because treatment-naïve disease encountered at first-line is more drug sensitive, the efficacy of new drugs in R/R disease has not been formally used to predict the efficacy of first-line treatments. To bridge this gap, we used the pop-TK model to simulate the selective pressure of first-line treatment and thereby to relate drug efficacy in R/R disease with first-line combination therapy.

A virtual cohort of R/R patients was created by simulating an RCHOP-treated cohort and selecting patients who experience disease progression (**Figure 4A**; **Figure S5**). This process represents the most drug-sensitive lymphomas being cured at first-line and therefore absent from R/R cohorts, as well as R/R lymphomas becoming even more drug resistant from the selective pressure of first-line chemotherapy. We next simulated subsequent therapies in this R/R cohort, fitting drug efficacy parameters (mean and inter-patient variance) to match observed PFS distributions of nine drug trials in R/R disease (44–52) (**Figure 4B**, column 1) and were consistent between trials for drugs tested in multiple R/R combinations (**Figure S6**). These parameters for drug efficacies were used to simulate the various first-line combination therapy trials, modeling the specific of each regimen in terms of whether a new drug was added to RCHOP (ibrutinib; lenalidomide; bevacizumab; venetoclax; tucidinostat), administered as maintenance after RCHOP (everolimus; enzastaurin), or replaced a drug in RCHOP (obinutuzumab replaced rituximab; polatuzumab-vedotin replaced vincristine) (**Figure 4B**, column 2; **Supplemental Table S2**). Predicted PFS hazard ratios (HR) fell within the 95% confidence intervals of observed HRs for eight out of nine trials (**Figure 4C**). The model overestimated the efficacy of RCHOP plus bevacizumab; in actuality this trial terminated early due to a high incidence of congestive heart failure, which also necessitated administering only half of the intended cumulative bevacizumab dose (25). This underscores that our model does not attempt to predict adverse events, a distinct but critical determinant of outcomes, although pop-TK models predicted the anti-tumor efficacy of each tolerable regimen.

**Figure 3.**
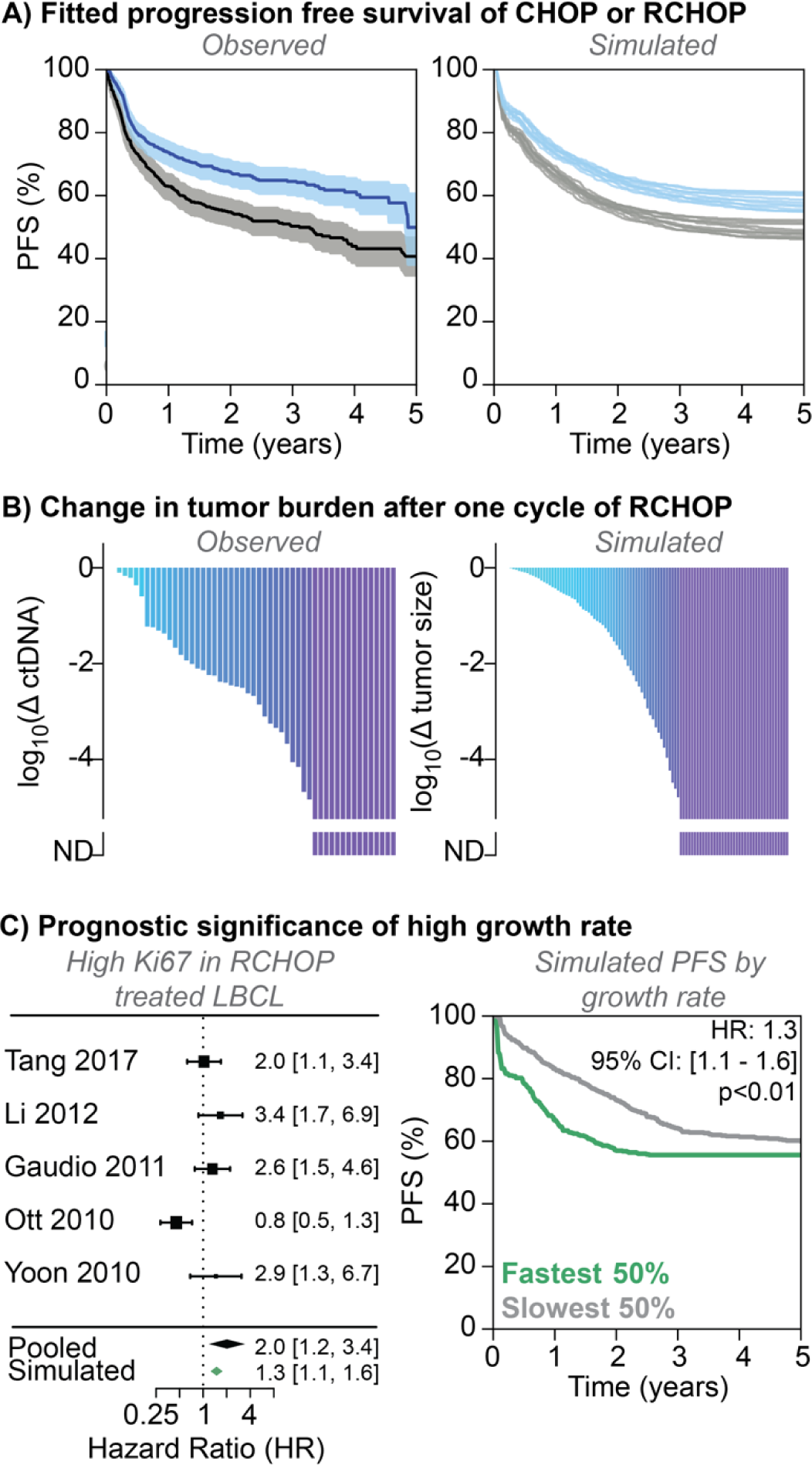
Model reproduces clinical efficacy of combination chemoimmunotherapy in DLBCL. (A) Progression free survival curves from a simulation of treatment with either six or eight cycles of either CHOP or RCHOP for an ensemble of parameter sets (right) compared to corresponding curves from the RICOVER-60 clinical trial (left, presented with 95% confidence intervals). **(B)** Waterfall plots of the simulated fold-change in tumor burden after one simulated cycle of RCHOP treatment (right) compared to the observed fold-change in clinically measured ctDNA after one cycle of RCHOP treatment (left) **(C)** Pooled Hazard Ratio (HR) analysis of the prognostic effect of high growth rate quantified by Ki67 in trials treating DLBCL patients with RCHOP compared to the simulated HR in the model (left). Simulated progression free survival curves for treatment with RCHOP stratified by growth rate (right).

**Figure 4.**
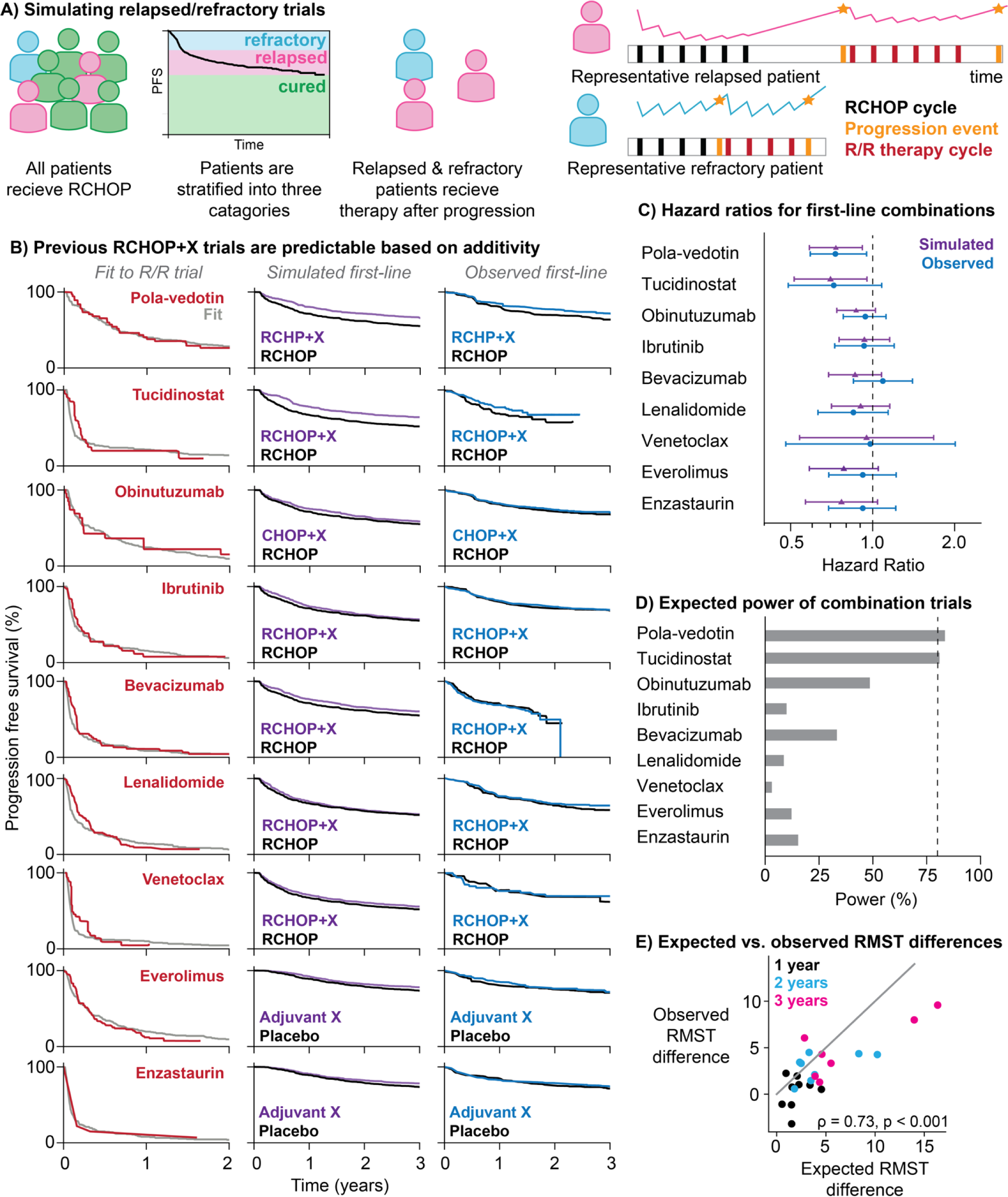
Model predicts efficacy of combination therapies at first line from drug efficacy of drugs in relapsed/refractory (R/R) DLBCL. (A) R/R trials are modeled by treating a cohort of simulated DLBCL patients with RCHOP, and then administering the second-line drug to patients who relapse or are refractory at the time of their progression event. **(B)** This approach to modeling R/R therapy has used to fit the model to R/R trial results of nine drugs: Polatuzumab-Vedotin, Tucidinostat, Obinutuzumab, Ibrutinib, Bevacizumab, Lenalidomide, Venetoclax, Everolimus,and Enzastaurin. The simulated R/R treatment (gray) was fit to the observed R/R progression free survival curves for each drug (red). The estimated efficacy of each drug was then used to simulate what would happen if that drug were added to the first line combination (purple), and the PFS distribution of this combination was compared to the simulated standard RCHOP treatment (black). The observed phase 3 combination clinical trial results are shown for the experimental (blue) and control (black) arms. **(C)** Comparison of the expected hazard ratio from the simulation (purple triangles) and the observed hazard ratio of the phase 3 trial (blue circles). **(D)** Expected statistical power to detect a significant positive result was calculated from the simulated combination trial using the actual design of the phase 3 clinical trial. Power is reported as the proportion of 1000 simulated trials that had a statistically significant positive result (p < 0.05 and 95% CI of HR < 1). The vertical dashed line represents 80% power. **(E)** The difference in restricted mean survival time (RMST) between the experimental and control arms was calculated for each trial simulated trial (x axis) and clinical trial (y axis). RMST was calculated at three time points (1, 2, and 3 years, denoted by color). The Spearman correlation between the simulated and clinical RMSTs was 0.73 (p < 0.001). Note that bevacizumab only had sufficient follow up to calculate the clinical RMST at one year.

We further evaluated clinical trial predictions by their expected statistical power, and by restricted mean survival time (RMST) for PFS, which quantifies area under the survival curve up to a defined time, providing an absolute measure of benefit. Each trial’s predicted HR and actual sample size was used to compute ‘statistical power’, being the expected likelihood of observing significantly improved PFS (*P*<0.05). The only regimens whose trial reached the conventional target of 80% power were R-CHP plus polatuzumab-vedotin (Pola-R-CHP; for higher risk DLBCL defined by IPI≥2) and RCHOP plus tucidinostat (for higher risk DLBCL defined by double expression of MYC and BCL2), which were indeed the only regimens to significantly improve PFS or EFS over RCHOP (**Figure 4D**) (24,44). Notably, our preliminary prediction of Pola-R-CHP versus R-CHOP was posted online before this trial’s results were first presented (**Figure S6**). Finally, predicted and observed changes in RMST at one, two, and three years were significantly correlated (**Figure 4E**; Spearman’s r = 0.73; *P*<0.001). These results show that the pop-TK model can use new drug efficacy in R/R clinical trials to predict the efficacy of new first-line combination therapies, including strategies of adding a drug, replacing a drug, or adjuvant therapy.

### Model-driven clinical trial design considerations

Having found that the pop-TK model explains recent trial results in DLBCL, we next used it to explore how future trials of combination therapy can improve patient outcomes. First, we aimed to understand why adding new drugs that do possess monotherapy activity failed to improve PFS in seven of nine clinical trials. Virtual trials of increasing numbers of drugs (*N* versus *N*+1 drugs), where each has the same average monotherapy efficacy, demonstrated diminishing returns; not simply in absolute survival change, but in relative risk reduction (**Figure 5A**). This suggests that improving survival beyond a 5-drug standard-of-care will either require new drugs to be dramatically superior to existing ones, or additional strategies such as synergistic combinations or use of biomarkers to develop precision drug combinations. We next investigated these possibilities using virtual trials.

**Figure 5.**
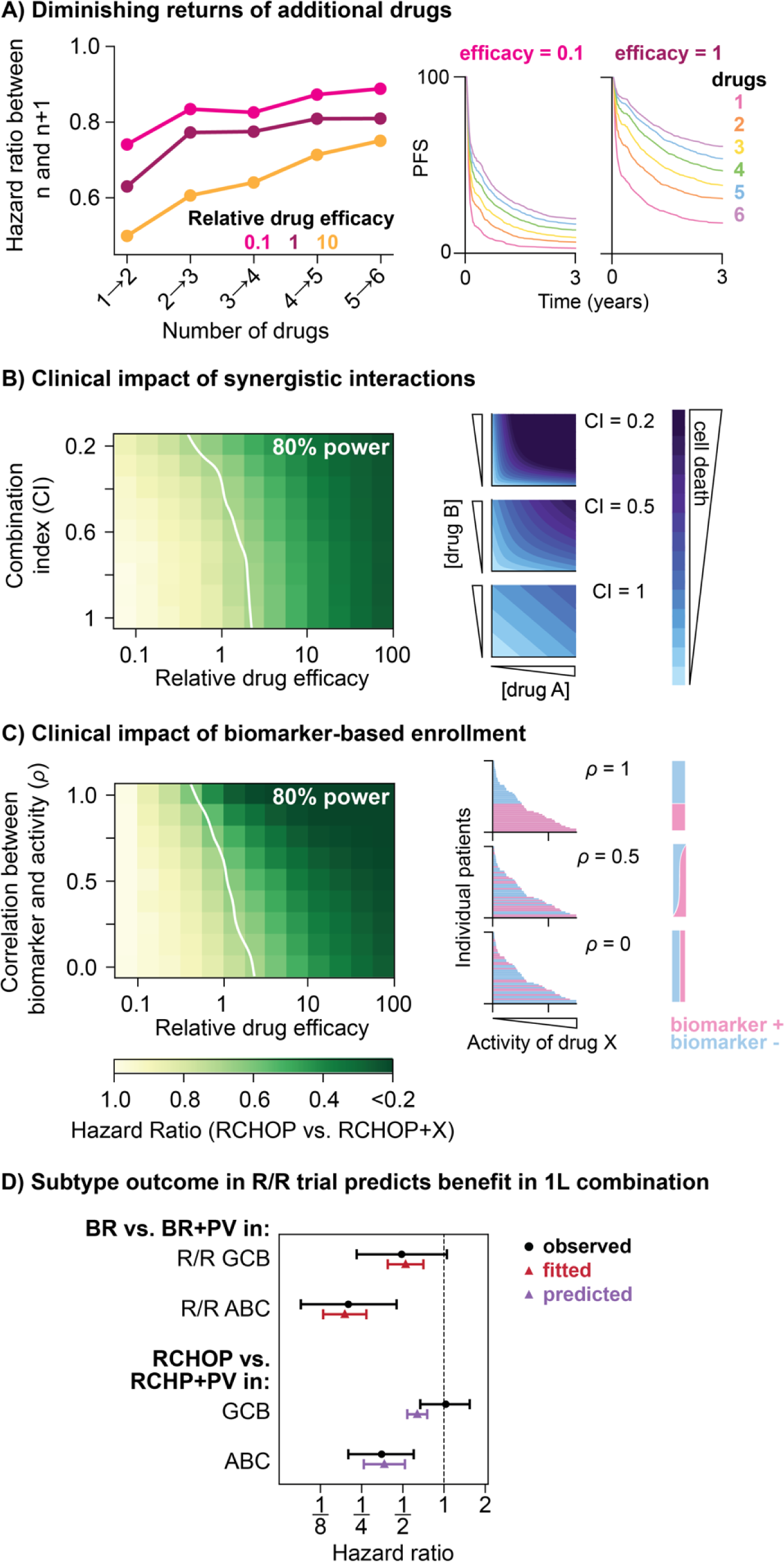
Impact of drug properties on clinical outcomes. (A) The hazard ratio of adding a new, equally effective, drug to a combination increases as the number of drugs. Examples PFS distributions are shown for drugs with relative efficacies of 0.1 or 1 (as effective as the drugs in CHOP). **(B)** Heatmap of HR of adding a drug with a given level of efficacy (x-axis) and a given level of synergistic interaction between the new drug and one of the drugs in RCHOP (y-axis). Efficacy is quantified as relative drug dose, where a dose of one represents a drug that is as effective as a single drug in the CHOP combination. Synergy is quantified as combination index (CI) where additive drugs have a CI of one and CI decreases with increasing synergy. Representative simulated isobolograms a shown for three levels of synergy. **(C)** Heatmap of HR of adding a drug with a given efficacy (x-axis) and a given correlation between biomarker and drug activity (y-axis). Representative stratification of patients into biomarker positive and biomarker negative populations for three levels of biomarker correlation (***ρ***). In both heatmaps, the white line indicates where the trials reach 80% power with a trial size of 400 patients per arm. **(D)** We applied the biomarker performance parameter to DLBCL subtype (Germinal Center B-Cell, GCB, or Activated B-Cell, ABC) as biomarker for Polatuzumab-Vedotin (Pola) activity. The correlation between DLBCL subtype and Pola activity was calibrated to the hazard ratios for Pola in R/R trials (red triangles). This correlation was then used to predict the subtype-specific hazard ratios for the RCHP-PV vs. RCHOP trial (purple triangles). Corresponding clinically observed hazard ratios are shown by black circles.

Unlike our previous models of non-interacting drug combinations (5–8), pop-TK models can simulate drug-drug interactions and their impact on clinical outcomes. We simulated combination therapy trials spanning a range of values for individual efficacy of a new drug, and a range of synergies between the new drug and a drug in RCHOP, from additive (combination index 1) to strong synergy (combination index 0.2, meaning 5-fold enhanced potency) (**Figure 5B**). Synergies of up to 2-fold potentiation had a negligible effect on trial results, but synergies greater than 2-fold in all patients had the potential to improve outcomes, provided the new drug’s single-agent activity was at least as good as drugs in the standard-of-care (i.e. RCHOP). These results suggest that enthusiasm for synergy in pre-clinical studies would be best limited to interactions with greater than two-fold enhanced potency (combination index <0.5), consistency across a majority of tumor models (e.g., panels of cell cultures), and involving drugs with substantial single-agent efficacy in humans.

Drugs with moderate overall response rates could also elicit greater activity in combinations if biomarkers are available to select patients most responsive to the added drug. We simulated combination therapy trials with biomarker-based enrollment, investigating the influence of overall drug efficacy and biomarker accuracy. Specifically, we defined a metric for the level of correlation between a biomarker and drug activity wherein a correlation of 1 indicates that a biomarker perfectly identifies the subset of patients who are most responsive to a new drug, and a correlation of 0 means the biomarker is not correlated with drug activity and using this biomarker for enrollment is no better than random patient selection (**Methods**). Virtual trials found that increasing biomarker performance led to superior hazard ratios and statistical power at all levels of drug efficacy (**Figure 5C**). A correlation of 0.5 between biomarker and drug activity, which leads to assignment of two-thirds of patients to the optimal treatment (**Figure S7**), was equivalent to a drug being 2.5-fold more potent overall, suggesting that biomarkers of moderate accuracy have substantial potential for precision combination therapy (**Figure 5C**).

As a tangible application, we analyzed biomarker performance for DLBCL cell of origin as a predictor of benefit from polatuzumab-vedotin (Pola). Many clinical trials have observed that DLBCL of Activated B Cell (ABC) origin is more responsive to Pola than Germinal Center B cell (GCB) origin (53,54), including a randomized phase 2 study of bendamustine and rituximab (BR) with or without Pola (47). We used a simulated trial of BR±Pola to calibrate the biomarker performance of subtype for Pola activity and found that 70% performance reproduced the hazard ratio difference between ABC and GCB subtypes (**Figure 5D**). Next, we used this biomarker performance to predict subtype specific hazard ratios for the phase 3 trial of Pola-R-CHP versus R-CHOP (24), and found that the large difference was predictable from the subtype-specific efficacy in the preceding phase 2 trial (**Figure 5D**). This example demonstrates the potential of early-phase biomarker observations to anticipate biomarker-based endpoints in phase 3 trials, to select for treatment those patients who derive the greatest survival benefits from new combination therapies.

## DISCUSSION

By modeling tumor heterogeneity as distributions of drug sensitivity, simulations of multi-drug response in patient populations can reproduce clinical trial results and predict the efficacy of new combination regimens. The presented model has three central assumptions: (1) the effects of drugs depend on dose, (2) drug sensitivity varies among the cancer cells within a patient, and (3) the average drug sensitivity of cancers varies between patients. We model these phenomena by mathematical approximations, but in essence these phenomena are indisputably real: cancers are not homogeneous. It is therefore important to note that our study has not proposed new mechanisms but sought to assemble established facts into a lifelike simulation of tumor responses across human populations. Adopting the mixed-effects approach of population-pharmacokinetics allows this ‘population tumor kinetics’ model to implicitly incorporate many complex aspects of tumor biology, such as how genetics, epigenetics, and microenvironment affect drug sensitivity, without needing to explicitly represent their mechanisms. This sidesteps many unsolved mechanistic questions but offers practical advantages for model fitting and application to designing regimens and trials.

Application of the pop-TK model to combination therapy for DLBCL explains and predicts important clinical outcomes with relatively few fitted parameters. When calibrated to PFS distributions of over 1000 patients treated with either CHOP or RCHOP (**Figure 3A**), the pop-TK model reproduced tumor shrinkage kinetics (**Figure 4B**; **Figure S4**) and the prognostic impact of growth rate (**Figure 3C**). By fitting a model of R/R patients to trials of new therapies in R/R patients, the pop-TK model was able to accurately predict the results of adding new therapies to first-line combinations (**Figure 4**). Virtual trials were also used to calculated the statistical power of real trials, not based on an aspirational hazard ratio, but using a data-based prediction of hazard ratio, and correctly predicted that only RCHP-polatuzumab-vedotin and RCHOP-tucidinostat were likely to be significantly superior to RCHOP (**Figure 4D**). Our model does not reproduce the incidence of relapses later than 3 years, but recent genomic studies have found that ‘late relapses’ of DLBCL usually result from cancer precursor cells acquiring new mutations to develop into related yet *de novo* cases of DLBCL (55). Thus, the inability of our model to reproduce late relapses supports the theory that late relapses are effectively new and chemotherapy-naïve lymphomas.

This model focuses on modeling tumor response and does not attempt to predict when or how combination toxicity occurs. When combinations result in levels of toxicity that require major adjustments to treatment, as occurred for RCHOP plus bevacizumab, modeling the intended trial design cannot be expected to reproduce actual outcomes (**Figure 4C**). However, this model could be used to understand the clinical impact of known treatment adjustments, including interruptions to treatment cycles or reduction of administered dose. Since the model does not predict what dose adjustments will be needed, we suggest that empiric data on incidence and magnitude of dose adjustments from phase 1 or 2 studies could be incorporated into models, to forecast whether those adjustments will compromise the anti-tumor efficacy of a new regimen.

This mechanistic model of combination therapy in DLBCL illuminated why some new drug combinations improve survival and others do not. The influence of three key factors were investigated, but we note that all are predicated on tolerability. First, a new drug’s monotherapy efficacy had the largest impact on the benefit of adding it to a combination. Therapies that improved first-line outcomes also conferred durable complete responses on a sizable proportion of patients with R/R disease. Second, using biomarkers to select patients whose disease is most likely to respond to a new drug could improve trial outcomes for the biomarker-positive subpopulation (**Figure 5C**). For example, pola-vedotin had a higher efficacy in R/R patients with ABC DLBCL, which predicted the observation in previously untreated DLBCL that the overall benefit of Pola-R-CHP derives from the ABC subtype (HR = 0.35) and not the GCB subtype (HR = 1.03) (24) (**Figure 5D**). This result suggests that cell-of-origin subtype should be considered when determining which patients receive pola-vedotin for previously untreated DLBCL (54). Third, synergistic drug interactions demonstrated limited ability to improve combination therapy except when strong synergy is exhibited by therapies that also possess substantial single-agent efficacy. This finding cautions against enthusiasm for synergy when it has modest magnitude, is spuriously present across pre-clinical models, or occurs among individually ineffective agents. Together, these clinical trial simulations suggest that developing new individually effective drugs and biomarkers for the most responsive disease subsets has the greatest potential to improve the success of first-line therapy for DLBCL.

In the future this model could be adapted to other therapies and types of cancer, by adjusting growth rates, dose response functions, and sensitivity distributions based on data from other diseases. We also anticipate that further biological details, such as spatial aspects of solid tumors, could be built atop the current framework. Simulations of clinical trials do not replace clinical trials, any more than simulations of airplanes replace airplanes, but in both cases, models can inform new designs and prevent failures. Reducing the frequency of unsuccessful phase 3 clinical trials has immense potential to lower the cost of developing superior cancer treatments, and accelerate improvement.

## METHODS

### Population Tumor Kinetics (pop-TK) model

The general form of pop-TK model is given by equation 1. This form includes flexibility for both the distribution of sensitivity phenotypes and dose response functions. In DLBCL both theory and experimentally observed drug sensitivity distributions support a log-normal sensitivity distribution (**Figure 1D-E**, **Figure S1**). The model includes cell-level cross-resistance between drugs, implemented as a correlation in drug sensitivities in a multivariate Gaussian distribution which has the following probability density function for *n* drugs:

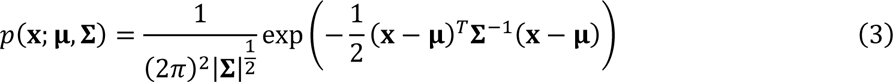

where **μ** is the is a 1 x *n* matrix of mean sensitivities, and **Σ** is the *n*x *n* variance matrix given by

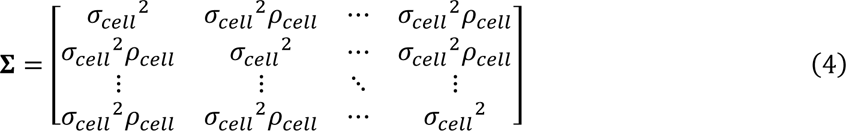

where *σ_cell_* is the standard deviation of the distribution of sensitivities, i.e., the level of heterogeneity between tumor cells, and *ρ_cell_* is the correlation between any two drugs in the combination, i.e., the extent of cross-resistance between drugs in a population of tumor cells.

In DLBCL experimental dose response measurements support an exponential dose response function (**Figure 1F**). Thus, the full pop-TK model equation which returns the number of cells surviving *c* cycles of *d* doses of *n* drugs is given by the multidimensional integral:

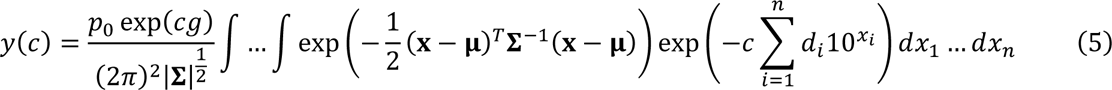

Computationally, multidimensional integration is solved using PyCuba (56). The code used for all simulations presented in this work is available in the linked Code Ocean capsule.

### Simulating pharmacological interactions

In the case of additive (non-interacting) drugs, the response to *n* drugs with doses d_1_, d_2_, … d_n_ is:

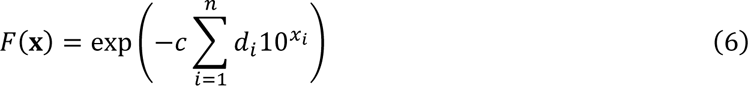

Pharmacological interactions (synergy or antagonism) are implemented by modifying the dose response to include drug-drug interaction terms. If we omit sensitivity distributions for explanatory purposes, response to drug pair with interaction *I* can be written:

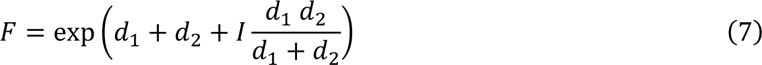

Where *I*>0 represents synergy, *I*<0 represents antagonism, and *I*=0 represents additivity, or no interaction. A general expression accounting for pairwise interactions *I_ij_* between drugs *i* and *j* is:

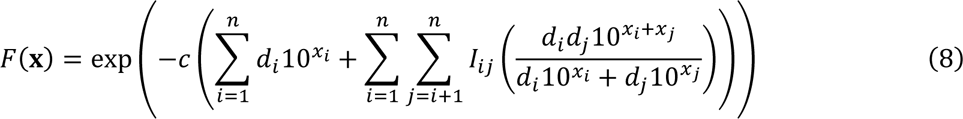

Interaction terms are related to ‘Combination Index’:

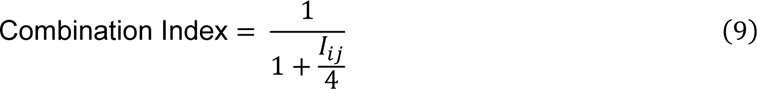

### Sampling virtual patient cohorts

Clinical trials were simulated as virtual patient cohorts, where each patient’s drug sensitivities, tumor growth rate, and initial tumor population are sampled from distributions as described below.

<u>Drug sensitivity:</u> Patients’ drug sensitivities were sampled from a multivariate log-normal distribution, with a level of correlation that describes cross-resistance between drugs. Specifically, a patient’s 1 x *n* drug sensitivity vector **μ** is sampled from a multivariate log-normal distribution: 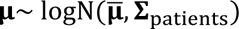 where **μ̄**-is mean sensitivity among all patients, and **Σ**_patients_ is

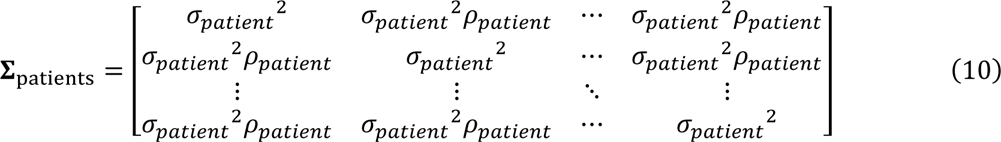

with *σ_patient_* being standard deviation of drug sensitivities among patients, and *ρ_patient_* being correlation in drug sensitivities (cross-resistance) among patients.

<u>Growth rate</u>: Each patient’s tumor growth rate *g* is sampled from a log-normal distribution with mean *μ_growth_* and standard deviation *σ_growth_*. A maximum growth rate *G* is imposed which corresponds to population doubling in 25 hours, which is the average doubling time of Burkitt lymphoma, the fastest growing human tumor (57).

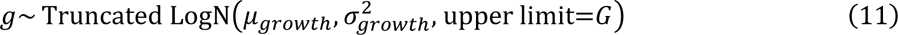

<u>Tumor burden:</u> The initial tumor burden for patient *n*, *p_o,n_*, depends on the patient’s tumor stage. This distribution was informed by clinical measurements of initial total metabolic tumor volume (TMTV) in patient cohorts stratified by stages I to IV (36). We generated log-normal distributions for each stages from using a common standard deviation within each stage (*σ_stages_*), a mean logarithm of tumor burden of stage IV disease (*μ_stageIV_*), and a difference in log (initial tumor burden) between stages (Δ_stages_) (**Figure S8**):

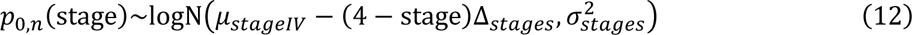

### Generating simulated progression free survival (PFS) distributions

A PFS distribution was generated by simulating the pop-TK model for each patient in a cohort. Progression times were computed from tumor growth rate and population at the end of treatment:

1. If the tumor population was greater than its initial size, progression was defined by the time required for the tumor to reach twice its initial population.
2. If the tumor population was reduced but remained more than one cell, progression was defined by the time required to regrow to its initial population.
3. If the tumor population was reduced to less than 1 cell, a patient was considered potentially cured, with a probability inversely proportional to residual tumor cell count; e.g. a patient with “0.1 cells” has 90% likelihood to be cured.

Cured patients were treated as censoring events at the longest follow-up time of the trial. Kaplan Meier curves were generated from a cohort of simulated progression and censoring events.

### Hazard ratio and power calculations

Hazard ratios were calculated using the Cox Proportional Hazards Model in the Lifelines python package (58). Expected statistical power of trials was calculated by simulating 1000 trials with the same design, sample size, and distribution of censoring events as the corresponding real trial. Power was the percentage of simulated trials that were ‘successful’ defined by Hazard Ratio significantly less than 1, at *P*<0.05.

### Model parameters

The model has 12 parameters, some of which are determined directly from experimental or clinical data, and some of which were fitted to reproduce PFS distributions (**Supplemental Table S1**).

<u>Initial tumor sizes (3 parameters):</u> Parameters defining the distribution of initial tumor burden were fit to clinical measurements of total metabolic tumor volume (TMTV) for a cohort of patients with varying Ann Arbor stage. This distribution is defined by three parameters: the standard deviation of tumor size for patients with the same Ann Arbor stage (*σ_stages_*), a mean tumor burden of stage IV cancer patients (*μ_stageIV_*), and the difference in initial tumor burden between stages (Δ_stages_) (equation 9) (**Figure S8**). For each simulation of a specific real trial, the frequency of patients with stage I to IV disease was matched to that reported in the real trial.

<u>Tumor growth rates (3 parameters):</u> Tumor growth rates *g* were sampled from a log-normal distribution with mean growth rate *μ_growth_*, standard deviation *σ_growth_*, and maximum growth rate *G* (equation 8). Average growth rate was estimated using the Metropolis Hastings algorithm. Standard deviation was determined from clinical measurements of growth rates of non-Hodgkin lymphomas (35). Maximum growth rate was set to the average doubling time of Burkitt lymphoma, the fastest growing human tumor (57).

<u>Drug sensitivity (6 parameters):</u> Drug sensitivity distributions for RCHOP were defined by six parameters: average sensitivity to the drugs comprising CHOP (*μ_CHOP_),* average sensitivity to Rituximab (*μ_R_*), intra-tumor heterogeneity in drug sensitivity (*σ_cell_*; applied to each drug), patient-to-patient heterogeneity in drug sensitivity (*σ_patient_*; applied to each drug), cross-resistance between drugs at the patient scale (*ρ_patient_*), and cross-resistance between drugs at the cellular scale (*ρ_cell_*). These parameters were fitted, via the Metropolis Hastings algorithm, to reproduce PFS distributions of the RICOVER-60 trial. Parameters *μ*, *σ_patient_*, *σ_cell_* each uniquely affect the shape of the PFS distribution (**Figure S2**). Because trials prior to the establishment of CHOP pre-date the modern histological definition of DLBCL, data is not available to estimate *μ* specific to each single agent in CHOP, and so a shared average sensitivity *μ_CHOP_* is used for these drugs.

### Metropolis-Hastings parameter estimation

Parameter values were fitted to reproduce the PFS distributions produced by 6 or 8 cycles of CHOP, with two week cycles, in the RICOVER-60 trial (59). A preliminary search screened 3 values each for 6 parameters (**Table S1**). 20 of these parameter sets produced PFS approximately near the efficacy of CHOP (residual errors being twice the 99% confidence interval of the clinical data; **Figure S3**). These parameter sets were used as starting values for Metropolis-Hastings random walks (60) by the procedure below.

1. Initiate the Metropolis-Hastings algorithm with one of the 20 candidate parameter sets.
2. For a specified chain length (500) perform the following steps:

a. Modify one randomly selected parameter from the previous step (*p*) by adding a normally distributed random value centered at zero with the parameter’s standard deviation from the prior distribution in the preliminary screen. The resulting parameter set is *p*’.
b. Check that the modified parameter is within the predetermined range. If it is outside of the range, set it to the closest value allowed. The resulting parameter set is *p*’.
c. Use the score function, *s*, to calculate the error value for the parameter set *p*’.
d. With the acceptance ratio *α* = *s*(*p*’)/*s*(*p*) accept or reject the parameter set *p*’. If accept, then *p*’ becomes *p*.

The Metropolis-Hastings algorithm was run 6 times for each of 20 starting parameter sets. In sum this tested 60,000 parameter sets (6 × 20 × 500). Pairwise distributions of all parameter sets in the 85% and 97.5% confidence intervals of CHOP’s PFS distribution are shown in **Figure S3**. Among 10 parameter sets that most closely reproduce the efficacy of CHOP, one set was closest to the centroid of these most plausible sets in every pairwise view, and was selected for further modeling. Using this selected parameter set, we estimated the average sensitivity to rituximab (*μ_R_*) that best reproduced the RCHOP arms of RICOVER-60 (59).

### Simulating and fitting trials in relapsed/refractory (R/R) cohorts

The R/R patient cohort was generated using a ‘6 drug’ model wherein all patients received first-line treatment with ‘RCHOP+X’ with ‘drug X’ having a dose of zero. Virtual patients who progressed during or after initial treatment (red and blue patients in **Figure 3A**) were selected to simulate second-line therapy with ‘drug X’ in R/R patients. Dosing schedules in virtual R/R trials were matched to schedules for each real trial of interest (**Table S2**). R/R trials were simulated across a grid of two parameters– dose of drug X (equivalent to adjusting tumor sensitivity to drug X) and inter-patient variance in sensitivity (*σ_patient_*). For each real trial of a ‘drug X’, we selected the parameter set that most closely fit clinically observed PFS distribution of that drug in R/R patients, by mean squared error in PFS over the reported duration of the trial, up to 2 years. These parameters defining the activity of each ‘drug X’ were used to simulate phase 3 trials of RCHOP+X with dose schedules matching each particular trial (**Figure 4B**; **Table S2**). Pola-vedotin had been trialed in combination with rituxutimab (47) and in combination with rituximab and bendamustine (61), and therefore we fit parameters for pola-vedotin to both of these trials, treating pola-vedotin as a more potent form of vincristine (O in RCHOP) because both vincristine and the cytotoxic payload of pola-vedotin, monomethyl auristatin E, are microtubule destabilizing agents that bind to *TUBB* (**Figure S6**).

### Simulating adjuvant therapy trials

Trials of adding Enzastaurin or Everolimus to RCHOP administered these agents as adjuvant therapy to patients with a complete response to RCHOP (26,27), and their primary endpoints included only patients with a complete response to RCHOP. Correspondingly, we modelled these trials as enrolling only patients with a complete response to RCHOP, defined as attaining a minimum tumor size of less than 1% of initial tumor burden. Time to progression was defined as the time from the end of RCHOP until the tumor reached its pretreatment size.

### Simulating biomarker stratified trials

Predictive biomarkers of drug response were simulated as a quantitative variable for each of *n* patients which is correlated with drug sensitivity, as described by:

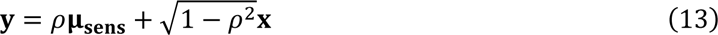

where **y** is a 1 x *n* matrix of biomarker level, **μ_sens_** is an 1 x *n* matrix of patient’s drug sensitivities with standard deviation *σ_patient_*, and **x** is a 1 x *n* matrix of normally distributed random values with mean 0 and standard deviation *σ_patient_*. A specified number of patients with the highest simulated biomarker levels are then enrolled on the trial (**Figure S7**).

### Assessing cell-to-cell drug sensitivity distribution

Cell-to-cell variation in sensitivity to single drugs in RCHOP was previously measured in DLBCL cultures by high complexity clone tracing and by genome-wide CRISPR inhibition screens under treatment (4). These data were re-analyzed to graph drug sensitivity distributions in Figure 1, showing log(enrichment) from 10^6^ barcoded DLBCL clones treated with each drug in RCHOP, and log(enrichment) of CRISPR inhibition guide RNAs (called ‘rho phenotype’) in DLBCL cultures treated with drugs in RCHOP (4).

## DISCLOSURES

A.C.P. has received consulting fees from AstraZeneca, Kymera, Merck, Novartis, and Sanofi, and research funding from Prelude Therapeutics. A.E.P. has received consulting fees from Respiratorious AB.

## Data Availability

All data used in the study are from previously published studies as cited in the manuscript.

## ACKNOWLEDGEMENTS

We thank the investigators and patients who participated in the clinical trials analyzed in this work. We thank Ash Alizadeh, Christopher Chidley, Haeun Hwangbo, Feng Fu, Jacob Pantazis, Sarah Patterson, Noah Schlachter, and Peter Sorger for discussions. A.E.P. is supported by NIGMS grant K12GM000678. This work is supported by NCI grant R01CA279968.

## Notes

### Funding Statement

The study was funded by NCI grant R01CA279968. A.E.P. is supported by NIGMS grant K12GM000678.

### Author Declarations

The study only used openly available de-identified human data from previously published clinical trials, available from the following links: RICOVER-60: https://pubmed.ncbi.nlm.nih.gov/18226581 Kurtz et al: https://pubmed.ncbi.nlm.nih.gov/30125215 R/R Enzastaurin: https://pubmed.ncbi.nlm.nih.gov/17389337 R/R Everolimus: https://pubmed.ncbi.nlm.nih.gov/21135857 R/R BR+polatazumab-vedotin: https://pubmed.ncbi.nlm.nih.gov/31693429 R/R R+polatuzumab-vedotin: https://pubmed.ncbi.nlm.nih.gov/30935953 R/R obinutuzumab: https://pubmed.ncbi.nlm.nih.gov/23835718 R/R bevacizumab: https://pubmed.ncbi.nlm.nih.gov/19373598 R/R venetoclax: https://pubmed.ncbi.nlm.nih.gov/28095146 R/R tucidinostat: https://pubmed.ncbi.nlm.nih.gov/34481935 Enzastaurin: https://pubmed.ncbi.nlm.nih.gov/27217449 Everolimus: https://pubmed.ncbi.nlm.nih.gov/29253068 polatuzumab-vedotin: https://pubmed.ncbi.nlm.nih.gov/34904799 obinutuzumab: https://pubmed.ncbi.nlm.nih.gov/28796588 bevacizumab: https://pubmed.ncbi.nlm.nih.gov/24895339 venetoclax: https://doi.org/10.1200/JCO.2024.42.16_suppl.7012 tucidinostat: https://doi.org/10.1200/JCO.2024.42.17_suppl.LBA7003

