## Supplementary Materials for "A mechanistic model of curative combination therapy explains lymphoma clinical trial results"

### Supplemental Figures

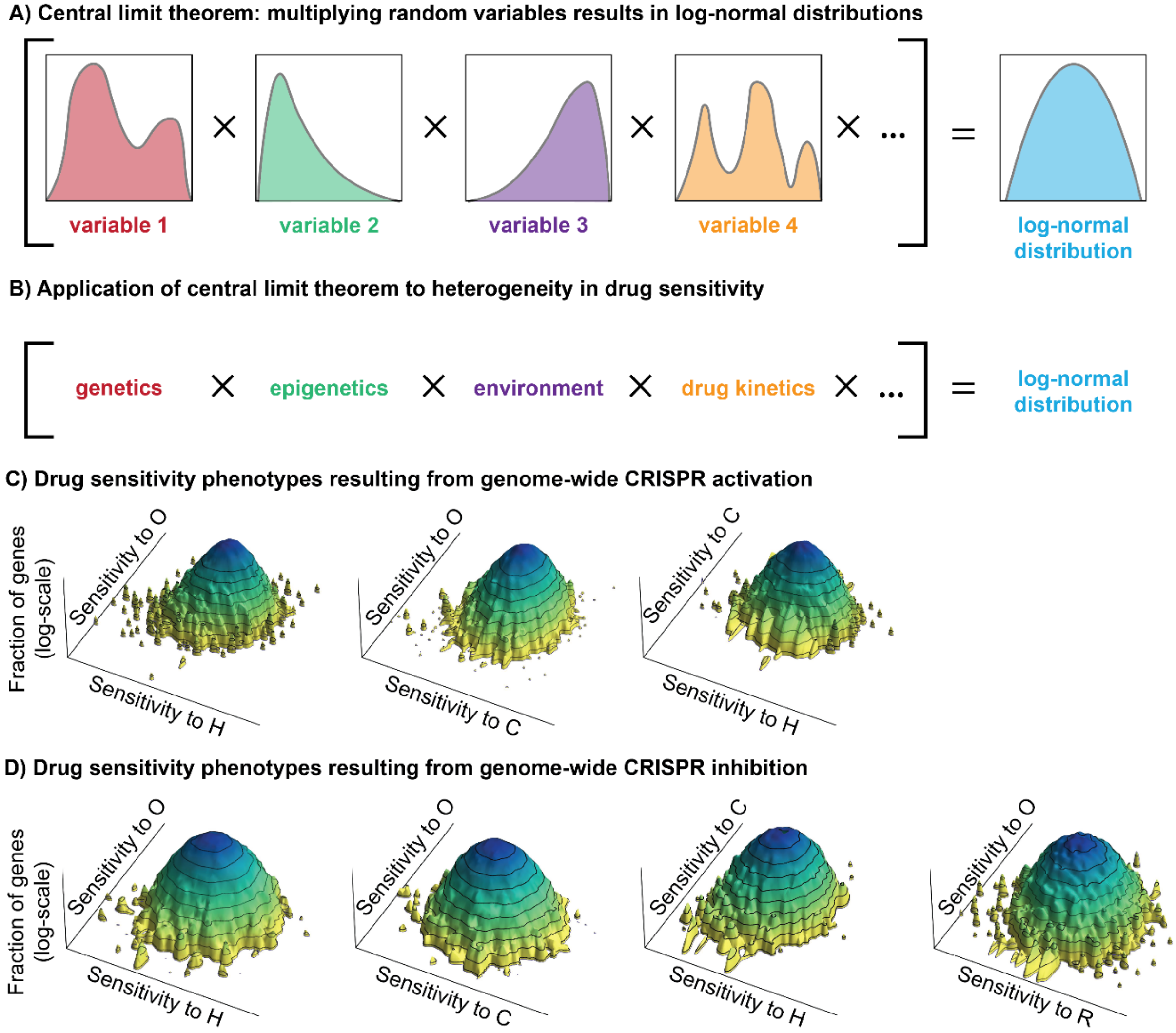


**Figure S1. Cell-to-cell variation in drug sensitivity is approximately log-normal. (A)** A consequence of the central limit theorem, when considered in the logarithmic domain, is that if a quantity is the product of many random variables, then the distribution of that quantity will be approximately log-normal, irrespective of the particular shapes of distributions of the underlying random variables, provided they have finite mean and variance, and that no single variable dominates the sum. **(B)** The central limit theorem can be applied to the distribution of tumor cells' sensitivity to a drug, where each variable is a biological factor that influences drug sensitivity, such as genetics, epigenetics, tumor environment, pharmacokinetics, and more. The consequence of this statistical principle is that highly complex tumor biology is likely to result in log-normal distributions of drug sensitivity. Reanalysis of data from **(C)** CRISPR activation (CRISPRa) and **(D)** CRISPR inhibition (CRISPRi) for each pairwise combination in the five-drug RCHOP combination demonstrates partly correlated log-normal distributions of drug sensitivity for DLBCL cells treated with these chemotherapies. Vertical axis is log (fraction of genes), and horizontal axes are 'rho phenotypes' for each drug, which is log-fold change in abundance of guide RNAs (average of 10 independent guide RNAs per target gene) between drug-treated and untreated control samples.

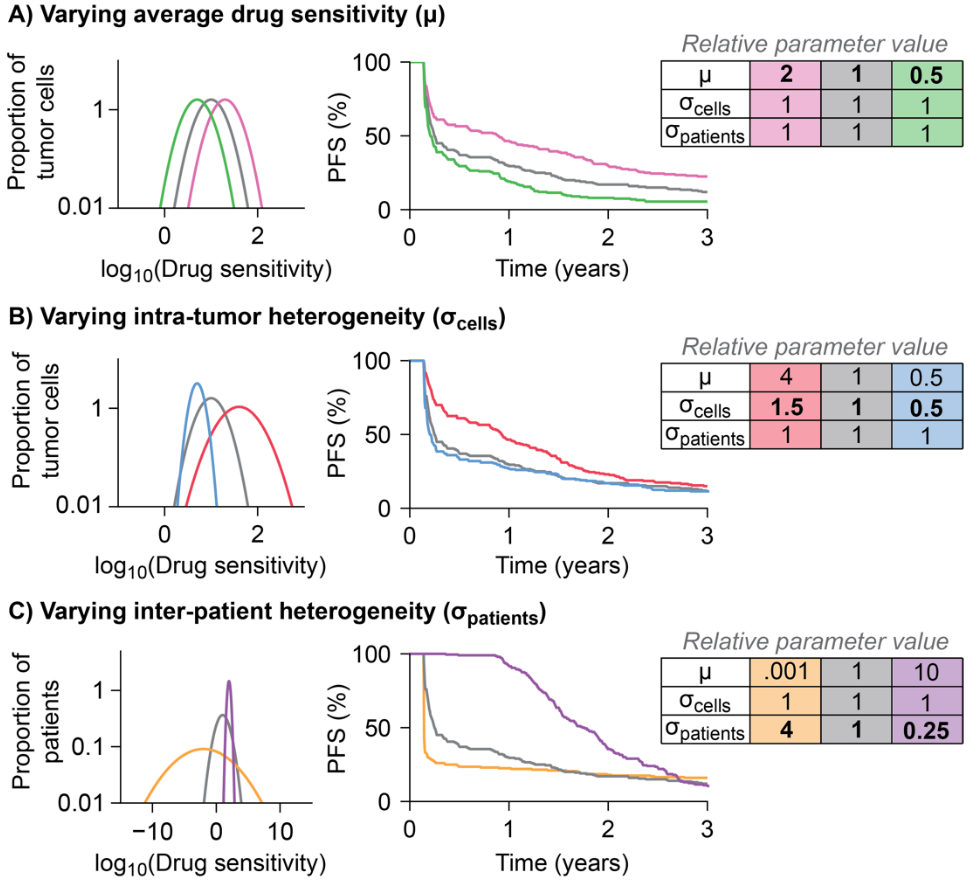


**Figure S2. Parameters affect the shape of PFS distributions in unique ways.** Three parameters that define drug sensitivity distributions were fit to clinically observed PFS distributions. These parameters cannot be directly measured in the clinic, but each parameter uniquely affects shapes of clinically measured PFS distributions, allowing them to be fitted to PFS. **(A)** First, the average drug sensitivity parameter (μ) for the distributions of both inter-patient and intra-tumor heterogeneity affects 3-year PFS without qualitatively changing the shape of the distribution. Thus, average drug sensitivity affects cure rate. **(B)** The standard deviation of the distribution of intra-tumor drug sensitivity ($\sigma_{cells}$) represents the extent of heterogeneity within patients. While adjusting μ to keep 3-year PFS constant, increasing $\sigma_{cells}$ leads to a higher proportion of patients having later relapses, changing the slope of the PFS distribution between years 1 and 3. **(C)** The standard deviation of the distribution of inter-patient drug sensitivity ($\sigma_{patients}$) represents the extent of heterogeneity between patients. While adjusting μ to keep 3-year PFS constant, increasing $\sigma_{patients}$ decreases the number of early progression events, and vice versa. Thus, the extent of patient-heterogeneity affects the slope of the PFS distribution in the first year. Note that each gray PFS distribution models treatment with one drug having the same parameters as individual drugs in RCHOP. Relative parameter values for alternative scenarios are shown in tables.

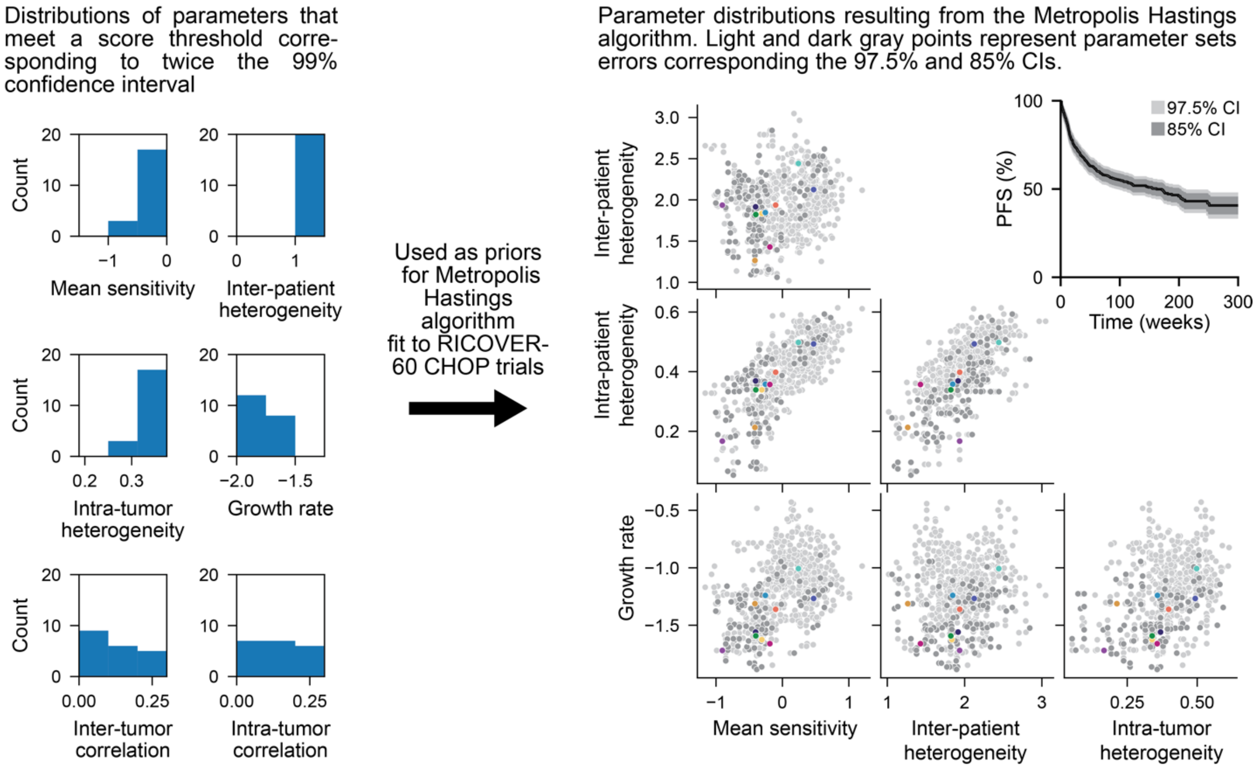


**Figure S3. Parameter estimation methods.** The first step of parameter estimation was an *in silico* screen of parameter sets by Monte Carlo sampling. The distribution of parameter sets that met the 99% confidence interval of the CHOP arms of the RICOVER-60 trials were used as priors for the Metropolis Hastings algorithm. The Metropolis Hastings algorithm identified many parameter sets whose modelled PFS fell within the 97.5% confidence interval of RICOVER-60 (light gray) and others within the 85% confidence interval (dark gray). From parameter sets in the 85% confidence interval, we selected 10 sets that collectively spanned the plausible parameter ranges (colored points) for subsequent analysis (**Figure S4**). Growth rate is shown on natural log scale. Drug sensitivity parameters (Mean, inter-patient heterogeneity, and intra-tumor heterogeneity) are shown on log10 scale, with heterogeneity parameters being standard deviation of a log-normal distribution.

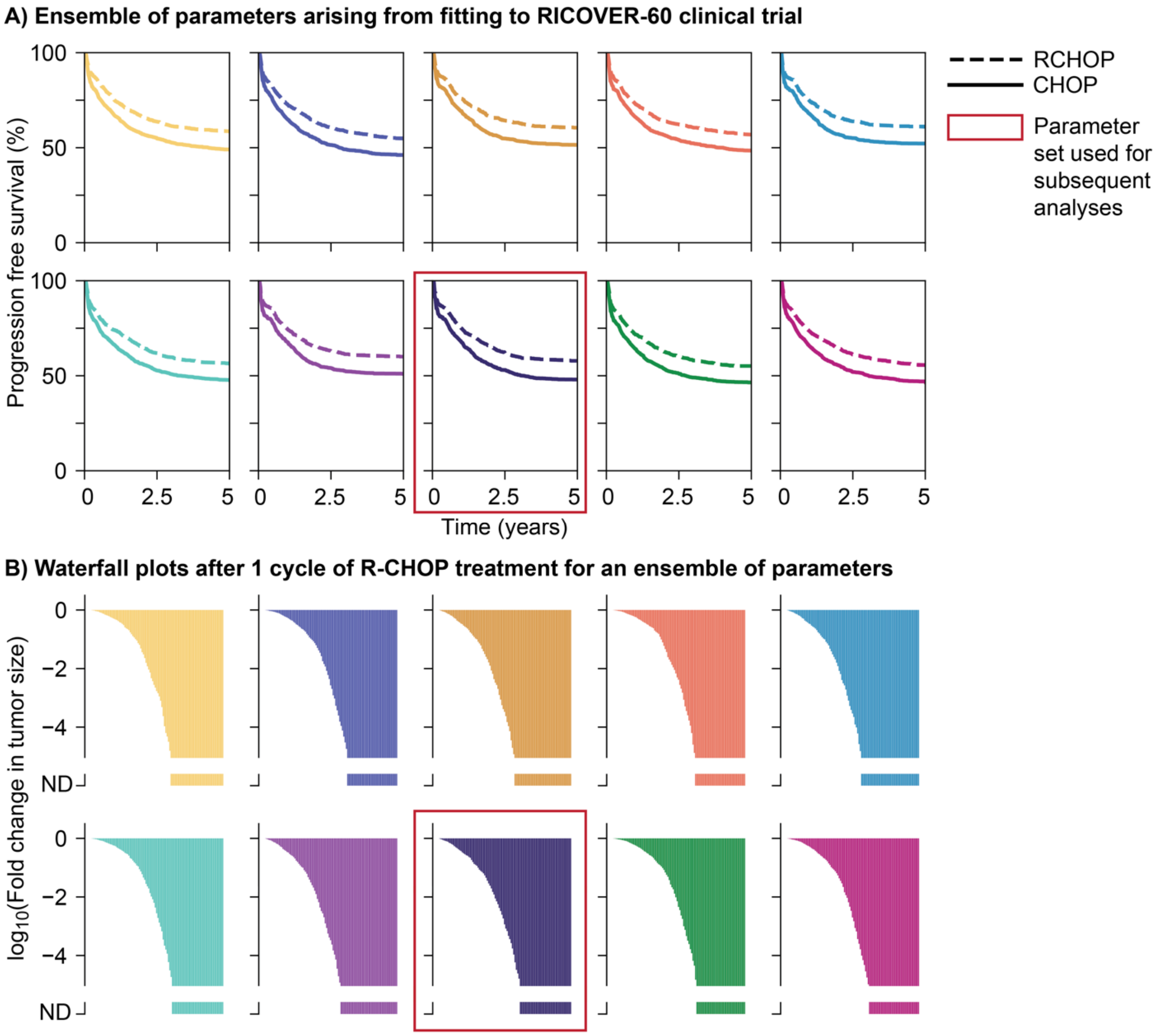


**Figure S4. PFS distributions and tumor size change distributions for each individual parameter set for the representative ensemble of parameters. (A)** PFS distributions for the response to CHOP (solid line) and RCHOP (dashed line) for each of 10 representative parameter sets. **(B)** Change in tumor population 'waterfall plots' for each parameter set. The parameter set used for subsequent analyses is outlined in red, and was closest to the centroid of plausible values in every scatterplot of parameter pairs in **Figure S3**. Colors for parameter sets are consistent across panels and Figure S3.

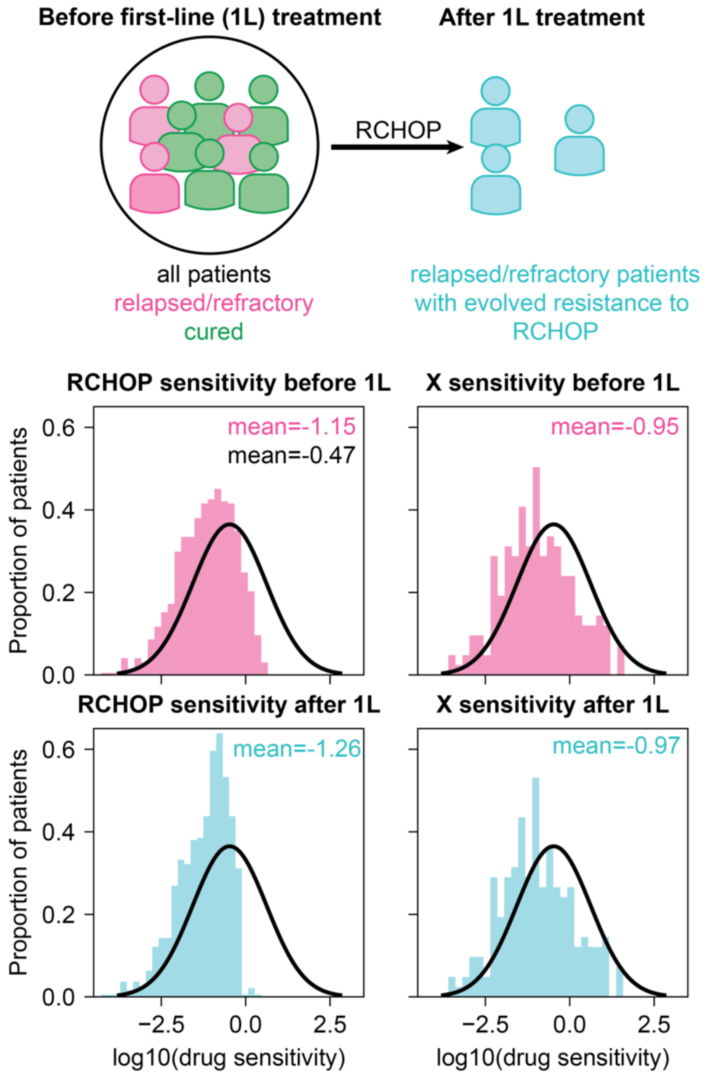


**Figure S5. Treatment with RCHOP decreases patients' sensitivities to RCHOP and novel agent (X).** Parameter estimation was used to define the sensitivity distribution of the entire pre-treatment population. Most patients are cured by RCHOP, but those patients who will not be cured (pink patients in top panel) will receive additional lines of therapy after RCHOP treatment fails. These patients are less sensitive to the drugs comprising RCHOP, since this standard treatment did not cure them, and their first-line of treatment results in the evolution of yet stronger drug resistance, as shown in **Figure 1C**. Distributions in pink show the pre-treatment drug sensitivity distributions of relapsed/refractory patients to either a drug with the same mechanism of action as a drug in RCHOP (e.g., rituximab, polatuzumab-vedotin, obinutuzumab) or a drug 'X' with novel mechanism of action. Distributions in blue show the post-treatment sensitivity distribution of relapsed/refactory patients. All distributions are compared to the pre-treatment drug sensitivity distribution of all patients (black).

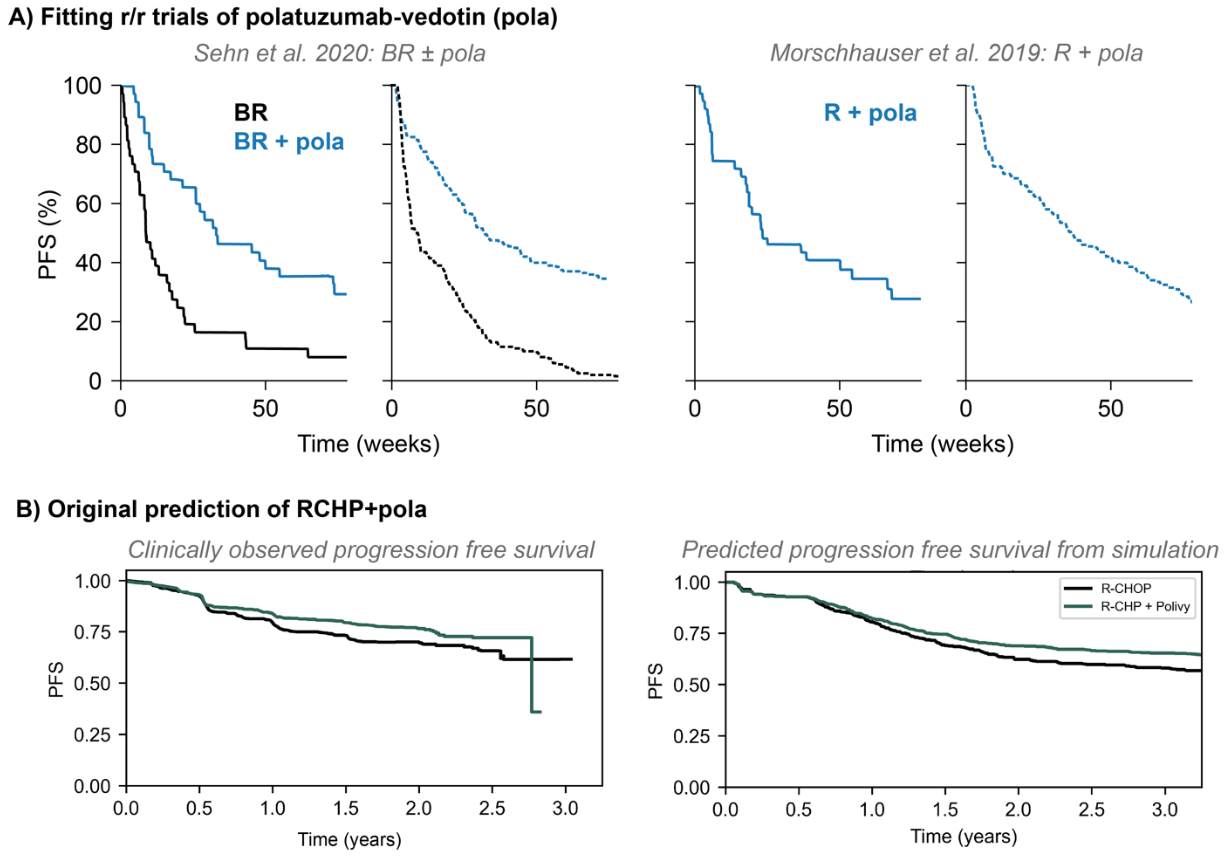


**Figure S6. Calibration to trials of pola-vedotin in R/R DLBCL and original POLARIX prediction.** (**A**) The parameters defining the relative acivity of pola were fit to both the trial comparing Bendamustine plus Rituximab (BR) with or without polatuzumab-vedotin (pola) and the trial of Rituximab plus pola (R-pola). Each drug in these combinations has an overlapping mechanism of action with a drug in RCHOP. Fitting the effective doses of pola-vedotin and bendamustine relative to their RCHOP counterparts to the R/R PFS curves resulted in the same parameter to define relative activity of pola compared to vincristine. In each case, the observed efficacy of pola is reproduced when modeling the administered dose as acting like a 10-fold higher dose of vincristine. (**B**) Initial prediction of a phase 3 trial comparing RCHOP to RCHP-pola predicted the results of the POLARIX trial on December 6, 2021, before the trial results were reported. Parameters and software implementing this early draft of the model varied slightly from the final model in this manuscript.

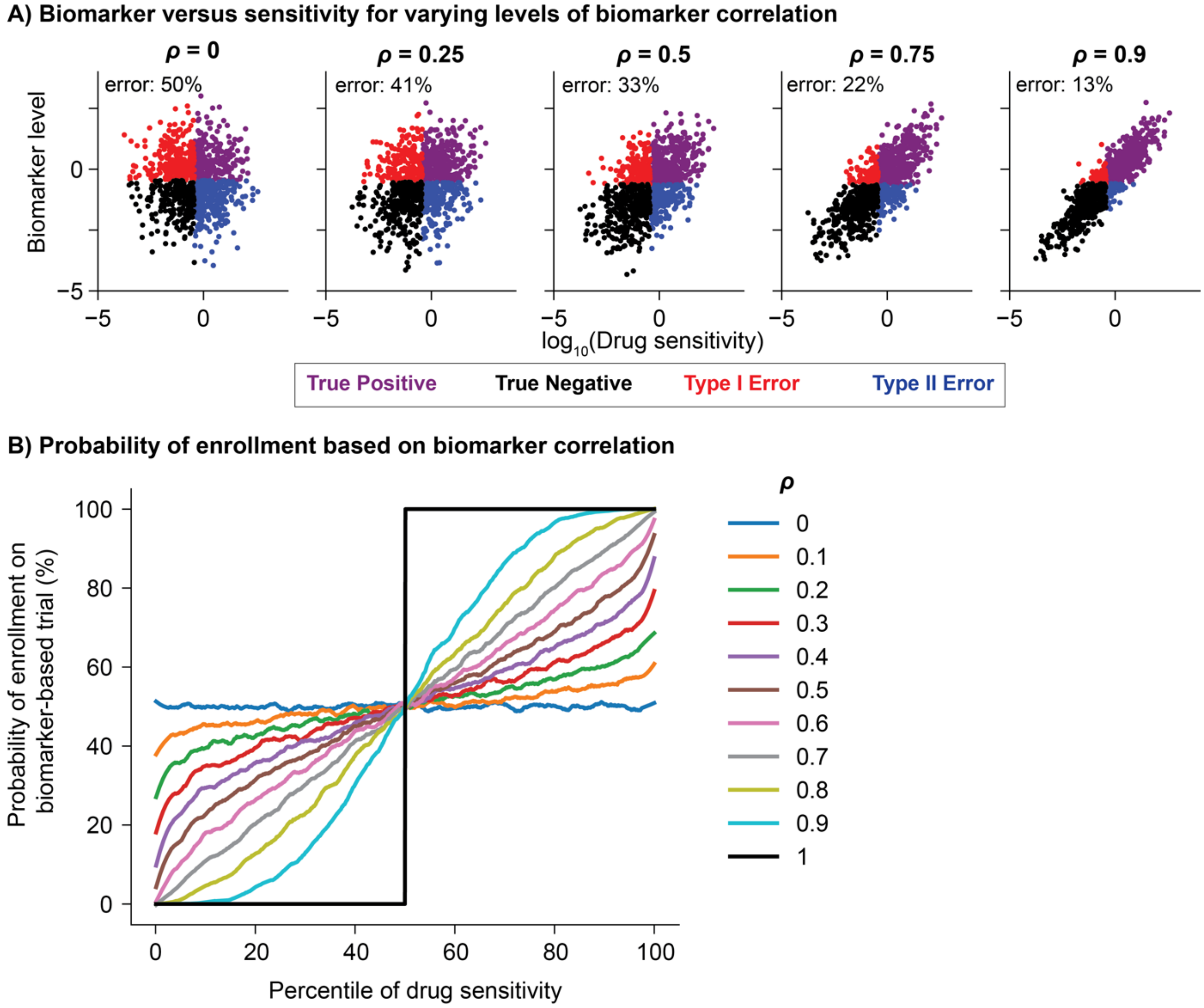


**Figure S7. Biomarker performance metric.** (**A**) A simulated biomarker level is calculated for every patient based on their drug sensitivity and the correlation between biomarker and sensitivity ($\rho$). For higher values of $\rho$ the biomarker level is more correlated with drug sensitivity and the error rate, either type I (false positive) or type II (false negative), decreases. (**B**) The plotted curves are for 500 simulations of a trial enrolling 50% of patients and show the probability of enrollment based on a patient's percentile of drug sensitivity. When $\rho$ is one the probability of enrolling the patients with the best responses is 100% and the probability of enrolling the patients with the worse responses is 0%. Conversely, when $\rho$ is zero there is a 50% probability of enrolling a patient regardless of their drug sensitivity. As $\rho$ increases the probability of enrolling a patient with high drug sensitivity increases.

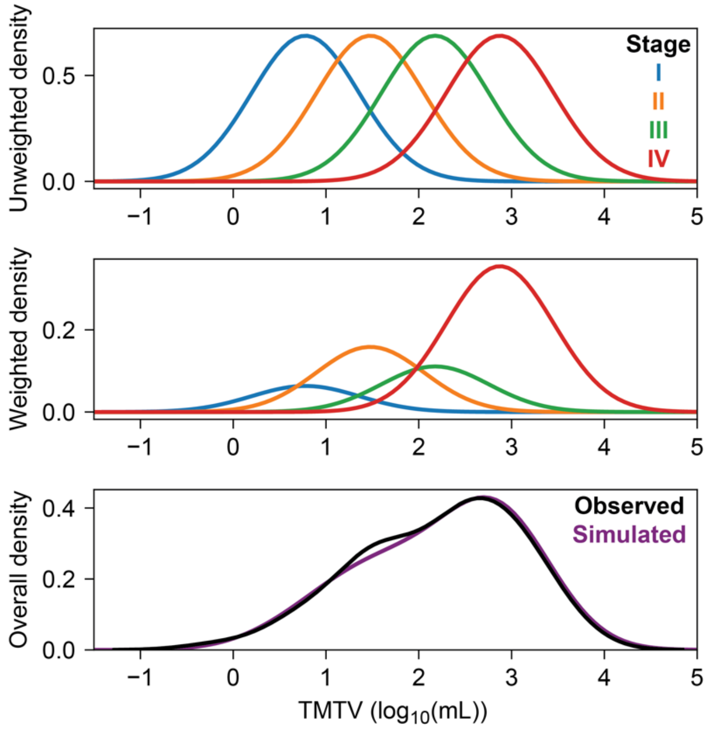


**Figure S8. Total metabolic tumor volume (TMTV) distribution by Ann Arbor Stage.** The top panel shows modeled distributions of tumor size for Ann Arbor stages I-IV. The middle panel shows the weighted density of these distributions based on the relative proportions of patients with each disease stage in Kurtz et al. 2018 (1). The bottom panel shows the cumulative density of all stages (purple) compared to the clinically observed distribution of TMTV (black).

### Supplemental Tables

| Distribution | Parameter | Description | Value | Units |
|---|---|---|---|---|
| $\log_{10}$(Drug sensitivity distributions) | $\mu_{sens,CHOP}$ | Average sensitivity to the drugs in CHOP | Estimated, allowed range (-∞, ∞), range of ensemble (-0.905, 0.47) | |
| | $\sigma_{inter}$ | Standard deviation of the patient sensitivity distribution (inter-patient heterogeneity) | Estimated, allowed range (0.01, ∞), range of ensemble (1.265, 2.442) | |
| | $\sigma_{intra}$ | Standard deviation of the tumor sensitivity distributions (intra-patient heterogeneity) | Estimated, allowed range (0.01, ∞), range of parameter ensemble (0.167, 0.498) | |
| | $\Delta\ \mu_{sens,R}$ | Average sensitivity to Rituximab | 2 – individually estimated | |
| | $\rho_{cell}$ | Correlation between drugs on the cellular level | Estimated, allowed range (0, 0.5), range of ensemble (0.001, 0.5) | none |
| | $\rho_{patient}$ | Correlation between drugs on the patient level | Estimated, allowed range (0, 0.5), range of ensemble (0.05, 0.431) | none |
| Growth rate distribution | $\mu_{growth}$ | Average growth rate | Estimated, allowed range (0, 2.0), range of ensemble (0.179, 0.365) | $\text{weeks}^{-1}$ |
| | $\sigma_{growth}$ | Standard deviation of the growth rate distribution | 0.615 – from clinical growth rate distributions (2) (Figure S6) | $\ln(\text{weeks}^{-1})$ |
| | $G$ | Maximum growth rate | 2.0 – fastest growing cancer (Burkitt's Lymphoma)(3) | $\text{weeks}^{-1}$ |
| Initial tumor burden distribution | $\mu_{stageIV}$ | Average tumor size of a phase IV tumor | $\mathbf{2.8751\times10^9}$ – from clinical data on TMTV distributions(1) (Figure S6) | cancer cells |
| | $\Delta_{stages}$ | Difference between average tumor size of two sequential phases | $\mathbf{7.0\times10^8}$ – from clinical data on TMTV distributions(1) (Figure S6) | cancer cells |
| | $\sigma_{stages}$ | Standard deviation of the tumor size distribution for a single stage | $\mathbf{5.8\times10^8}$ – from clinical data on TMTV distributions(1) (Figure S6) | cancer cells |

**Table S1. Parameter table.** List of all model parameters and how their values were determined.

| Drug | Drug target | Monotherapy trial | Mono. trial size | Mono. trial design – simulation design | Combo trial | HR for PFS vs. RCHOP | Combo. trial size | Combo. trial design | Dose of drug X |
|---|---|---|---|---|---|---|---|---|---|
| Bevacizumab | VEGF-A | Stropeck et al., 2009 (4) | 45 | 10 mg/kg every two weeks until progression<br>--<br>Dosed every two weeks for 1 year | MAIN (5) | 1.09 | RCHOP: 397<br>RCHOPX: 390 | Either 8 cycles of RCHOP-21 (80%) or 6 cycles of RCHOP-14 (20%) +X or +placebo. Continued X administered every 3 weeks for 12 months | 10/15 mg/kg |
| Enzastaurin | PKCβ | Robertson et al., 2007 (6) | 55 | 500 mg daily for up to 6 28-day cycles<br>--<br>Dosed every 28 days for 6 cycles | PRELUDE (7) | 0.92 | RCHOP: 254<br>RCHOPX: 504 | Patients received 6-8 cycles of RCHOP prior to enrollment. Patients either received 500 mg X or placebo daily for up to 3 years | 500 mg daily |
| Everolimus | mTORC1 | Barnes et al., 2013 (8) | 77 | 10 mg daily for up to 6 28-day cycles<br>--<br>Dosed every 28 days for 6 cycles | PILLAR-2 (9) | 0.92 | RCHOP: 370<br>RCHOPX: 372 | Patients had a complete response to RCHOP prior to enrollment. Patients received either 10 mg X or placebo daily for up to 1 year | 10 mg daily |
| Ibrutinib | BTK | Wilson et al., 2015 (10) | 38 ABC<br>20 GCB | 560 mg daily until progression or intolerable<br>--<br>Dosed every 21 days for 1 year | PHOENIX (11) | 0.93 | RCHOP: 419<br>RCHOPX: 419 | ABC patients received 6-8 21-day cycles of RCHOP + X or placebo | 560 mg daily |
| Lenalidomide | CRBN | Witzig et al., 2011 (12) | 108 DLBCL | 25 mg daily on days 1-21 of a 28-day cycle until progression or intolerable<br>--<br>Dosed every 28 days for 1 year | ROBUST (13) | 0.85 | RCHOP: 285<br>RCHOPX: 285 | Patients received 6 21-day cycles of RCHOP + X or placebo with X on days 1-14 of the cycle | 15 mg daily |
| Obinutuzumab | CD20 | Morschhauser et al., 2013 (GAUGUIN) (14) | 25 | 1,600 mg in the first 21-day cycle and 800 mg in subsequent 21-day cycles<br>--<br>Dosed at 2X for 1 cycle and then 1X every 21 days for 1 year (in R dimension) | GOYA (15) | 0.94 | RCHOP: 712<br>RCHOPX: 706 | Patients received 8 21-day cycles of RCHOP + X or placebo | 1000 mg |
| Polatuzumab-Vedotin | CD79B and β-tubulin | Sehn et al., 2020 (16) | BR+X: 40<br>BR: 40 | 90 mg/m$^2$ Bendamustine<br>375 mg/m$^2$ Rituximab<br>1.8 mg/kg Polatuzmab -vedotin<br>--<br>Dosed every 21 days for 6 cycles | POLARIX (17) | 0.73* | RCHOP: 439<br>RCHOPX: 440 | Patients received 8 21-day cycles of RCHOP + X or placebo | 1.8 mg/kg |
| Venetoclax | BCL2 | Davids et al., 2017 (18) | 34 | 1,200 mg Venetoclax daily until progression or intolerable<br>--<br>Dosed every 21 days for 1 year | Alliance 051701 (19) | 0.98 | RCHOP: 59<br>RCHOPX: 60 | Patients received 6 21-day cycles of RCHOP + X or placebo with X on days 1-5 of the cycle | 800 mg |
| Tucidinostat | HDAC | Sun et al., 2021 | 20 | 30 mg Tucidinostat twice weekly until progression or intolerable<br>--<br>Dosed every 21 days for 1 year | Zhao et al. | 0.72* | RCHOP: 212<br>RCHOPX: 211 | Patients received 6 21-day cycles of RCHOP + X or placebo followed by 6 additional cycles of X or placebo. X was administered twice weekly for the first two weeks of each cycle | 30 mg |

**Table S2. RCHOP+X trials.** *Indicates a significant improvement

| Description | Year | Arm 1 (Experimental) | Arm 2 (Control) | Location | N patients |
|---|---|---|---|---|---|
| RICOVER-60 6 cycles (20) | 2006 | EFS for 6 cycles RCHOP14 | EFS for 6 cycles CHOP14 | Figure 3A | 613 |
| RICOVER-60 8 cycles (20) | 2006 | EFS for 8 cycles RCHOP14 | EFS for 8 cycles CHOP14 | Figure 3A | 609 |
| TMTV distribution (1) | 2018 | Initial TMTV for each enrolled patient | -- | Figure S8 | 67 |
| ctDNA change distribution (1) | 2018 | Fold-change in ctDNA after 1-2 cycles of RCHOP | -- | Figure 3B | 50 |
| R/R trial of Enzastaurin (6) | 2007 | Enzastaurin monotherapy | -- | Figure 4B | 55 |
| R/R trial of Everolimus (8) | 2013 | Everolimus plus rituximab | -- | Figure 4B | 77 |
| R/R trial of Lenalidomide (12) | 2011 | Lenalidomide monotherapy | -- | Figure 4B | 108 |
| R/R trial of Ibrutinib (10) | 2015 | Ibrutinib monotherapy | -- | Figure 4B | 58 |
| R/R trial of BR ± pola (16) | 2020 | Bendamustine, rituximab, and pola-vedotin (BR+pola) | Bendamustine and rituximab (BR) | Figures 4B, S5A | 80 |
| R/R trial of R + pola (21) | 2019 | Pola-vedotin plus rituximab | -- | Figure S5A | 39 |
| R/R trial of Bevacizumab (4) | 2009 | Bevacizumab monotherapy | -- | Figure 4B | 45 |
| R/R trial of Obinutuzumab (14) | 2013 | Obinutuzumab monotherapy | -- | Figure 4B | 25 |
| R/R trial of Venetoclax (18) | 2017 | Venetoclax monotherapy | -- | Figure 4B | 34 |
| R/R trial of Tucidinostat (22) | 2021 | Tucidinostat monotherapy | -- | Figure 4B | 20 |
| Phase 3 Enzastaurin combo (7) | 2016 | Enzastaurin following complete response to RCHOP | Placebo following complete response to RCHOP | Figure 4B | 758 |
| Phase 3 Everolimus combo (9) | 2018 | Everolimus following complete response to RCHOP | Placebo following complete response to RCHOP | Figure 4B | 742 |
| Phase 3 Lenalidomide combo (13) | 2021 | 6 cycles Lenalidomide and RCHOP | 6 cycles placebo and RCHOP | Figure 4B | 570 |
| Phase 3 Ibrutinib combo (11) | 2019 | 8 cycles Ibrutinib and RCHOP | 8 cycles placebo and RCHOP | Figure 4B | 838 |
| Phase 3 Pola-vedotin combo (17) | 2021 | 8 cycles polatuzumab-vedotin and RCHP | 8 cycles RCHOP | Figures 4B, S5A | 879 |
| Phase 3 Bevacizumab combo (5) | 2014 | 8 cycles Bevacizumab and RCHOP | 8 cycles placebo and RCHOP | Figure 4B | 787 |
| Phase 3 Obinutuzumab combo (15) | 2017 | 8 cycles Obinutuzumab and CHOP | 8 cycles RCHOP | Figure 4B | 1,418 |
| Phase 2/3 Venetoclax combo | 2024 | 6 cycles of Venetoclax and RCHOP | 6 cycles of RCHOP | Figure 4B | |
| Phase 3 Tucidinostat combo | 2024 | 6 cycles of Tucidinostat and RCHOP + 6 cycles of Tuc. | 6 cycles of RCHOP | Figure 4B | |

**Table S3. All digitized data included in manuscript.** All data are in DLBCL patients, and all data are PFS unless otherwise noted. Data is publicly available in Code Ocean capsule.
